# A phase IV, multi-centre, randomized clinical trial comparing two pertussis-containing vaccines in pregnant women in England and vaccine responses in their infants

**DOI:** 10.1101/2021.01.21.21250225

**Authors:** Christine Elizabeth Jones, Anna Calvert, Jo Southern, Mary Matheson, Nick Andrews, Asma Khalil, Hannah Cuthbertson, Bassam Hallis, Anna England, Paul T Heath, Elizabeth Miller

**Affiliations:** Paediatric Infectious Diseases Research Group, St George’s, University of London, London, UK; St George’s University Hospitals NHS Foundation Trust, London, UK; Faculty of Medicine and Institute for Life Sciences, University of Southampton and NIHR Southampton Clinical Research Facility and NIHR Southampton Biomedical Research Centre, University Hospital Southampton NHS Foundation Trust; Immunisation and Countermeasures, National Infection Service, Public Health England, London, UK; National Infection Service, Public Health England, Porton, Salisbury, UK; Statistics, Modelling and Economics Department, Public Health England, London, UK

**Author notes:** Corresponding author: Christine E Jones, Clinical and Experimental Sciences, Room LF102, F Level, South Academic Block, University Hospital Southampton NHS Foundation Trust, Tremona Road, Southampton, SO16 6YD, E.

**Keywords:** Maternal vaccination, immunization, infant, immune, response, pregnancy, vaccine

## Abstract

**Background:** Pertussis vaccines containing three or five pertussis antigens are recommended in pregnancy in many countries, but no studies have compared the effect on infants’ antigen-specific immunoglobulin G (IgG) concentrations. The aim of this study was to compare anti-pertussis IgG responses following primary immunization in infants of mothers vaccinated with TdaP5-IPV (low dose diphtheria toxoid, tetanus toxoid, acellular pertussis [five antigens] and inactivated polio) or TdaP3-IPV in pregnancy (three pertussis antigens).

**Methods:** This multi-centre phase IV randomized clinical trial was conducted in a tertiary referral centre and primary care sites in England from 2014-2016. Women were randomized to receive TdaP5-IPV (n=77) or TdaP3-IPV (n=77) at 28-32 gestational weeks. A non-randomized control group of 44 women who had not received a pertussis-containing vaccine in pregnancy and their 47 infants were enrolled postpartum.

**Results:** Following infant primary immunization, there was no difference in the geometric mean concentrations (GMCs) of anti-pertussis toxin, filamentous haemagglutinin or pertactin IgG between infants born to women vaccinated with TdaP5-IPV (n=67) or TdaP3-IPV (n=63). However, the GMC of anti-pertussis toxin IgG was lower in infants born to TdaP5-IPV and TdaP3-IPV vaccinated mothers compared to infants born to unvaccinated mothers (n=45) (geometric mean ratio: 0.71 [0.56-0.90] and 0.78 [0.61-0.98], respectively); by 13 months of age, this difference was no longer observed.

**Conclusion:** Blunting of anti-pertussis toxin IgG response following primary immunization occurs in infants born to women vaccinated with TdaP_5_-IPV and TdaP_3_-IPV, with no difference between maternal vaccines. The blunting effect had resolved by 13 months of age. These results may be helpful for countries considering which pertussis-containing vaccine to recommend for use in pregnancy.

**Clinical Trials identifier:** ClinicalTrials.gov: NCT02145624

## Background

In 2012, an antenatal pertussis vaccination programme was introduced in the United Kingdom (UK) as an outbreak measure following a significant increase in cases of pertussis and of pertussis-related infant deaths (1). Two vaccines have been recommended for use in pregnancy in the UK programme – REPEVAX-IPV (2012-2014, 2020) and BOOSTRIX-IPV (2014 -2019) – with high vaccine effectiveness observed for both vaccines (2). It is assumed that this reflects the increased levels of maternally-derived pertussis-specific antibodies present in the first weeks of life in infants born to vaccinated mothers. However, high levels of maternal antibody can interfere with the infant’s response to primary immunization, a phenomenon known as blunting. This has been shown for five- and three-pertussis antigen component vaccines, but no study has directly compared these vaccines with respect to their impact on infant responses to primary immunization (3-8).

## Methods

The primary objective of this trial was to assess antibody responses to pertussis antigens following primary immunization in infants born to women who were randomized in pregnancy to receive one of two pertussis-containing vaccines and to compare these responses to those in infants born to unvaccinated women. Transplacental transfer and concentrations of pertussis-specific antibodies prior to primary vaccination and 13 months of age were also assessed.

### Study design

In this phase IV, multi-centre, randomized clinical trial, pregnant women were randomized to receive either REPEVAX-IPV or BOOSTRIX-IPV (Additional file 1: Protocol). A contemporaneous non-randomized control group of infants born to women who had not received a pertussis-containing vaccine in pregnancy were also recruited.

Pregnant women, aged 16-45 years at enrolment, receiving antenatal care at St George’s University Hospitals NHS Foundation Trust, or in primary care sites in Gloucestershire and Hertfordshire, were eligible to participate. Exclusion criteria included: a bleeding disorder, receipt of a pertussis-containing vaccine in the previous 12 months, receipt of a blood product within the preceding 3 months, any contraindication to vaccination specified in the “Green Book” Immunisation against Infectious Disease (9).

Following written, informed consent, pregnant women were randomized 1:1 to receive either REPEVAX-IPV (2 International Units [IU] diphtheria toxoid [DT], 20 IU tetanus toxoid [TT], 2.5 μg pertussis toxoid, 5 μg, filamentous haemagglutinin [FHA], 3 μg pertactin [PRN], 5 μg fimbriae [FIM] types 2 and 3 and inactivated poliovirus [IPV, 40 D-antigen unit Type 1, 8 D-antigen unit Type 2, 32 D-antigen unit Type 3]; TdaP_5_-IPV; Sanofi Pasteur) or BOOSTRIX-IPV (2 IU DT, 20 IU TT, 8 μg pertussis toxoid, 8 μg FHA, 2.5 μg PRN and IPV [40 D-antigen unit Type 1, 8 D-antigen unit Type 2, 32D-antigen unit Type 3]; TdaP_3_-IPV; GlaxoSmithKline (GSK)).

A computerised block randomisation list was generated by the study statistician, with sites allocated blocks of sequential numbers (block size 8).

Visits for pregnant women occurred at 28-32 weeks of gestation, up to 7 days post-partum, and at 13 months following delivery. Cord blood or, if not obtained, peripheral blood up to 7 days of age, was collected from infants born to vaccinated mothers. All infants had peripheral blood collected at 2, 5 and 13 months of age. The study was approved by the MHRA, NHS Health Research Authority and City & East Research Ethics Committee (14/LO/0141). The study was registered with ClinicalTrials.gov (NCT02145624) prior to study commencement.

### Intervention

Pregnant women received either TdaP_5_-IPV or TdaP_3_-IPV as a 0.5 mL intramuscular injection into the left upper arm at 28 – 32 weeks of gestation. Infants received routine vaccines according to the nationally recommended schedule at the time of the study, Table 1.

**Table 1:** The routine immunization schedule recommended in the UK during the study period

| Vaccine | Trade name and Manufacturer | Recommended age at vaccination (months) |
| --- | --- | --- |
| **DTaP<sub>3</sub>-IPV-Hib** | Infanrix-IPV-Hib, GSK | 2, 3, 4 |
| **Meningococcal serogroup B** | Bexsero, GSK | 2, 4, 12 |
| **13-valent pneumococcal conjugate** | Prevenar13, Pfizer | 2, 4, 12 |
| **Oral live attenuated rotavirus** | Rotarix, GSK | 2, 3 |
| **Meningococcal serogroup C conjugate** | NeisVac-C, GSK | 3 |
| **Hib-MenC** | Menitorix, GSK | 12 |
| **Measles-mumps-rubella (MMR)** | Priorix; Sanofi Pasteur | 12 |

Abbreviations: DTaP<sub>3</sub>-IPV: diphtheria, tetanus, acellular pertussis (three pertussis antigens, 25 µg each of pertussis toxoid and filamentous haemagglutinin and 8 µg of pertactin), inactivated poliovirus, *Haemophilus influenzae* type B; IPV: Inactivated poliovirus; Hib: *Haemophilus influenzae* type B; MenC: Meningococcal serogroup C; GSK: GlaxoSmithKlein

### Outcomes

The primary outcome was fold difference in anti-pertussis toxin (PT) immunoglobulin G (IgG) geometric mean concentration (GMC) in infants at 5 months of age whose mothers received TdaP_5_-IPV or TdaP_3_-IPV in pregnancy. Secondary outcomes included: placental transfer of IgG to pertussis antigens in infants born to vaccinated mothers and GMC in infants born to vaccinated and unvaccinated mothers at 2,5 and 13 months of age.

### Safety

Women were observed for 20 minutes post vaccination for any immediate reaction. Adverse events and serious adverse events (SAEs) were collected for women and infants at each study visit.

### Laboratory assays

Serum IgG to PT, FHA, PRN and FIM 2&3 were quantified using enzyme-linked immunosorbent assays (ELISAs), developed in-house and performed by staff blinded to group allocation. All assays have been validated in accordance with International Conference on Harmonisation guidelines and use the 1^st^ World Health Organisation (WHO) International Standard Pertussis Antiserum (human) 06/140 (NIBSC, Item No. 06/140). The lower limit of detection (LLOD) of the assays are: 2.128 IU/ml (PT), 0.715 IU/ml (FHA), 0.806 IU/ml (PRN), 0.636 U/ml (FIM 2&3) with results less than the LLOD, assigned a value half of the LLOD.

### Statistical methods

Sample size calculation was based on the standard deviation of the post-primary vaccination anti-PT IgG GMC of 0.28 IU/ml from a previous study, generated using the the same validated PT ELISA (10). A sample size of 65 per study arm enabled detection of 1.38 fold differences or greater between study arms with 80% power at a 5% significance level. To allow for loss to follow-up, the target sample size was 75-80 in each vaccinated group, with 50 mother-infant pairs in the non-randomized control group.

To increase power, the data for infants born to unvaccinated mothers was supplemented with data from 19 infants from another study conducted at the same sites, at a similar time for which laboratory analysis was performed by the same laboratory using the same assays (<u>NCT01896596</u>). Infants in this study received Infanrix hexa (GSK) instead of Infanrix-IPV-Hib at 2, 3 and 4 months and had been randomized to receive one of three different Men C vaccines at 3 months of age. Blood samples were collected at 5 and 13 months.

A modified intention to treat analysis was performed; a per protocol analysis was not performed as there were fewer than 10% of individuals with data that differed between these populations. Pertussis IgG GMCs and geometric mean fold ratios (GMR) between groups, with 95% confidence intervals (CI) were calculated (Additional materials 2: Statistical Analysis Plan). Normal errors regression on log-transformed data was used to investigate the effect of pre-primary antibody on post primary GMCs. Two-sided 5% significance was shown when the 95% CI for the GMR or fold effect of pre-primary antibody did not contain unity. The placental transfer ratio (PTR) was calculated as the geometric mean ratio of infant-to-mother specific IgG at delivery; normal errors regression on logged ratios with adjustment for the interval between vaccination and birth were calculated. The effect of interval from birth to blood test, sex, ethnicity, birthweight and previous maternal pertussis vaccination were also examined by multiple regression when assessing transfer ratios. The proportion of mothers and infants experiencing SAEs was calculated for each group. Missing data were excluded from analyses and analyses were performed using Stata version 13.

## Results

Between October 2014 and October 2015, 154 pregnant women were enrolled and randomized to receive either TdaP_5_-IPV (n=77) or TdaP_3_-IPV (n=77); 159 infants were born to these women, 144 were included in the study (Figure 1). Twenty-five women who had not received a pertussis-containing vaccine in pregnancy were recruited in the postnatal period, to whom 27 infants were born; data from an additional 19 infants of unvaccinated mothers were included from the Infanrix hexa study.

**Figure 1:**
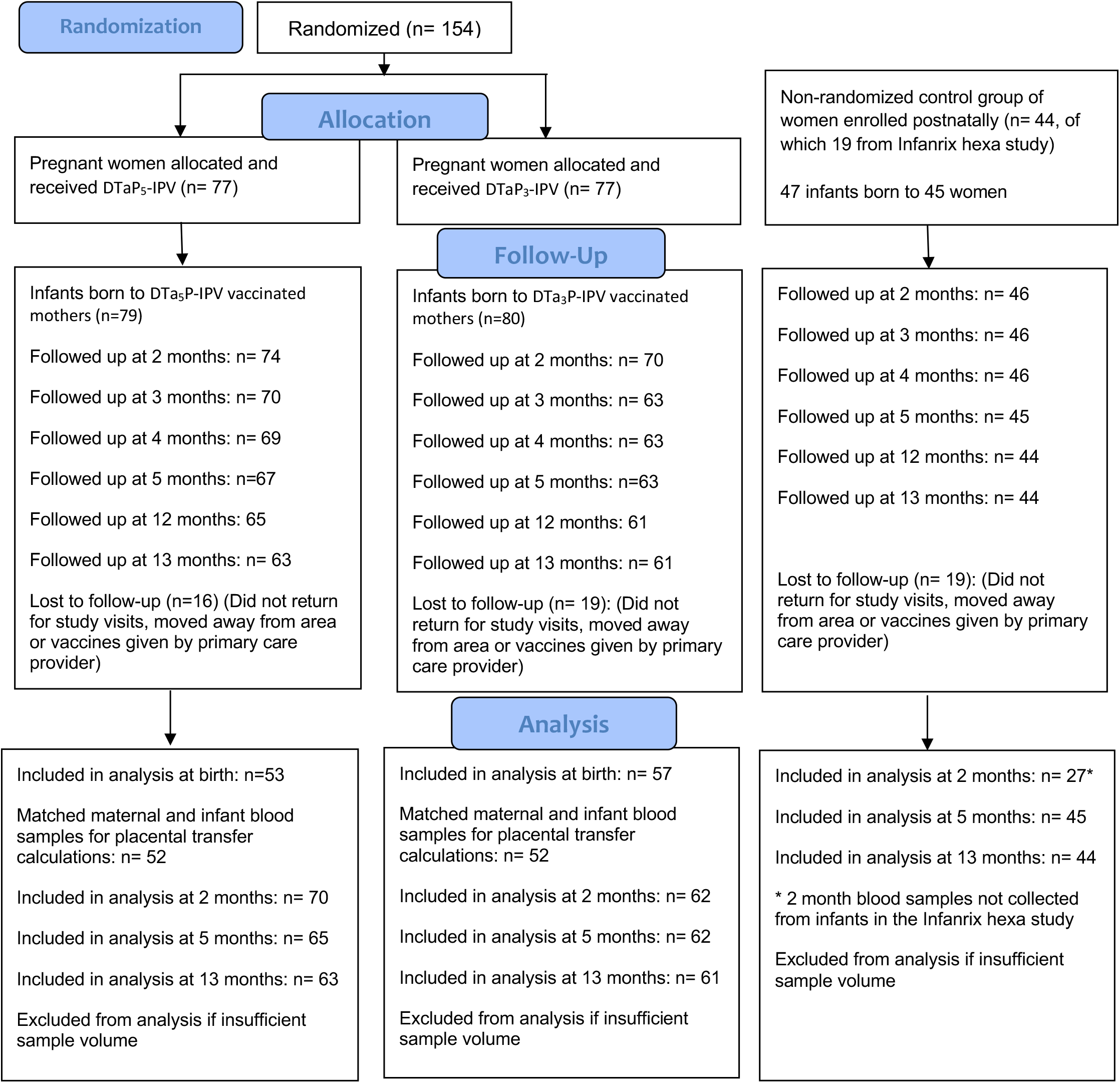
Consort flow diagram: Randomized groups of women receiving either TdaP_5_-IPV or TdaP_3_-IPV in pregnancy and their infants and a non-randomized group of women not receiving a pertussis-containing vaccine in pregnancy

Demographic and clinical characteristics of participating mother-infant pairs are described in Table 2. There were no baseline demographic differences between groups of women. [INSERT TABLE 2 – see end of manuscript for table]

**Table 2:** Baseline demographics of participating women and infants

| Characteristic | Maternal Tdap<sub>3</sub>-IPV<br>(n=77) | Maternal Tdap<sub>5</sub>-IPV<br>(n=77) | Unvaccinated<br>mothers<br>(n=25) | Unvaccinated mothers<br>from Infanrix hexa<br>study (n=18) |
| --- | --- | --- | --- | --- |
| **Women** |  |  |  |  |
| Age, years, median (range) | 34 (20-42) | 32 (21-44) | 31 (20-47) |  |
| **Ethnicity** |  |  |  |  |
| White | 63 | 63 | 17 | 16 |
| Asian | 6 | 7 | 2 | 1 |
| Black | 3 | 3 | 5 | 0 |
| Chinese | 1 | 1 | 0 | 0 |
| Mixed | 3 | 3 | 1 | 1 |
| Other | 1 | 0 | 0 | 0 |
| **Site** |  |  |  |  |
| Gloucestershire, UK | 3 | 5 | 0 | 5 |
| Hertfordshire, UK | 33 | 28 | 3 | 3 |
| South London, UK | 41 | 44 | 22 | 10 |
| Median gestation at vaccination, week (range) | 29 (28-34) | 29 (28-33) |  |  |
| Median interval from antenatal vaccination to delivery, weeks (range) | 10 (1-14) | 10 (3-14) |  |  |
| Median interval from delivery to blood sample post-delivery, days (range) [number of samples] | 3 (0-10) [51] | 2 (0-9) [57] |  |  |
| **Infants** |  |  |  |  |
|  | Maternal Tdap<sub>3</sub>-IPV<br>(n=70) | Maternal Tdap<sub>5</sub>-IPV<br>(n=74) | Unvaccinated<br>mothers<br>(n=27) | Unvaccinated mothers<br>from Infanrix hexa<br>study (n=19) |
| Twin | 6 | 4 | 4 | 2 |
| Female, n (%) | 33 (42%) | 38 (51%) | 13 (48%) | 10 (52%) |
| Median gestation at birth, weeks (range) | 39 (31-42) | 40 (33-42) | 38.5 (34-42) | 39 (37-42) |
| Median birth weight, kg (range) | 3.39 (1.54-4.69) | 3.52 (1.72-4.93) |  | 3.42 (2.74-4.20) |
| Cord blood collected, n | 20 | 21 |  |  |
| Infant peripheral blood collected, n | 37 | 32 |  |  |
| Median interval from birth to 1<sup>st</sup> infant blood sample, days (range) [number of samples] | 3 (0-10) [57] | 2 (0-9) [53] |  |  |
| Median age at pre-vaccination blood sample and vaccine dose 1, days (range)[number of samples] | 60 (53-87) [62] | 62 (50-80) [70] | 64 (54-83) [27] | 58 (54-81) [19] |
| Median age at vaccine dose 2, days (range) [number of samples] | 89 (81-113) [63] | 91 (81-108) [69] | 93 (82-125) [27] | 89 (82-121) [19] |
| Median age at vaccine dose 3, days (range) [number of samples] | 120 (108-154) [63] | 116.5 (110-143) [68] | 123 (110-154) [27] | 122 (112-156) [19] |
| Median interval to blood sample after vaccine dose 3, days (range) [number of samples] | 28 (21-47) [62] | 28 (21-55) [65] | 28 (21-52) [27] | 28 (21-40) [18] |
| Median age at “1 year” vaccination, days (range) [n vaccinated] | 372 (351-404) [61] | 372 (358-413) [65] | 373 (366-394) [27] | 377 (354-395) [17] |
| Median interval from “1 year” vaccination to blood sample, days (range) [number of samples] | 28 (21-48) [61] | 29 (21-42) [63] | 29 (21-52) [27] | 31 (21-51) [17] |

Abbreviations: TdaP<sub>3</sub>-IPV: low dose diphtheria, tetanus, acellular pertussis (three pertussis antigens), inactivated poliovirus TdaP<sub>5</sub>-IPV: low dose diphtheria, tetanus, acellular pertussis (five pertussis antigens), inactivated poliovirus; UK: United Kingdom
The following cells are empty as data was not collected from the women recruited in the postnatal period who had not received a pertussis-containing vaccine in pregnancy: maternal age (Infanrix hexa group only), gestation at vaccination, median interval from antenatal vaccination to delivery, median interval from delivery to blood sample post-delivery, birth weight, number of cord blood samples or peripheral blood at birth collected, median interval from birth to 1<sup>st</sup> infant blood sample.

### Placental transfer (Table 3)

The PTR of IgG was greater than 1 for all pertussis antigens, with no difference observed according to maternal vaccine received. There was a strong positive correlation between infant and maternal GMCs of IgG for PT, FHA, FIM 2&3 and PRN.

**Table 3:**
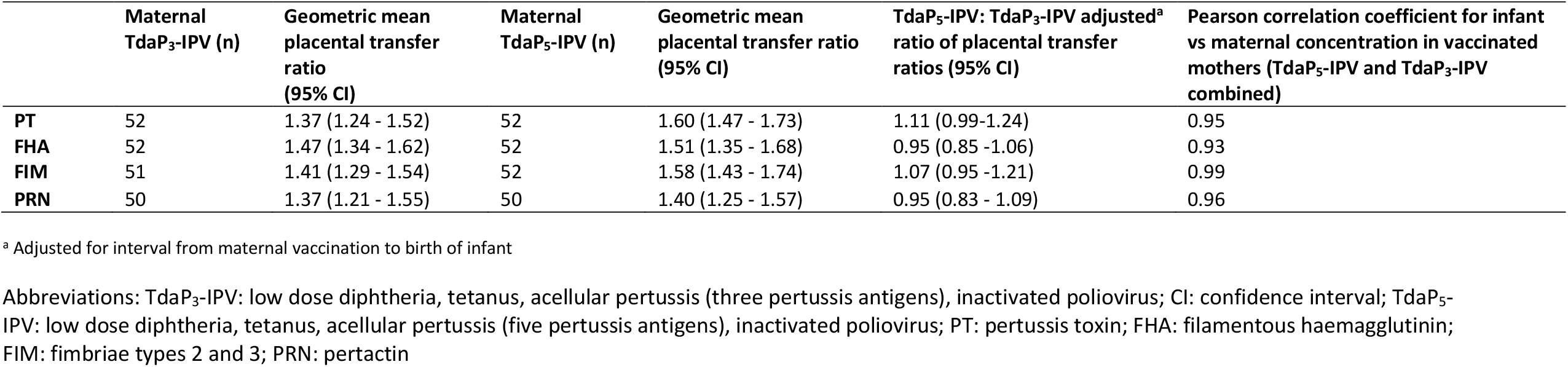
Placental transfer of pertussis-antigen specific IgG from TdaP_3_-IPV and TdaP_5_-IPV-vaccinated mothers to their infants

Multivariable analysis was performed to identify the factors associated with PTR. Time from vaccination to birth was significantly associated with PTR, with a fold change of 1.08 (95% CI 1.05-1.10), 1.10 (1.08-1.13), 1.06 (1.04-1.09) and 1.11 (1.08-1.14) for PT, FHA, FIM 2&3 and PRN respectively per week. If a cord sample was not collected, the time at which the infant peripheral blood sample was obtained was significantly associated with PTR with a fold change of 0.97 (0.96-0.99), 0.97 (0.95-0.99), 0.98 (0.96-1.00), 0.99 (0.96-1.01) for PT, FHA, FIM and PRN respectively per day from birth to blood sampling. Other factors assessed were not associated with the PTR.

### Infant antibody concentrations at 2 months of age (Table 4)

Prior to primary vaccination, infants born to mothers vaccinated with TdaP_5_-IPV had lower GMC of anti-PT and FHA IgG compared to infants born to TdaP_3_-IPV vaccinated mothers (GMR 0.64 [95% CI 0.43-0.94] and 0.48 [0.35-0.65] respectively). Infants born to mothers vaccinated with TdaP_5_-IPV had higher GMCs of anti-FIM IgG (GMR 8.71 [5.2-14.58]). Anti-PRN IgG were similar for infants born to TdaP_5_-IPV and TdaP_3_-IPV vaccinated mothers.

**Table 4:** Pertussis-antigen specific IgG GMC in infants at birth, 2, 5 and 13 months of age

| Antigen | Time- point | Maternal TdaP<sub>3</sub>-IPV |  | Maternal TdaP<sub>5</sub>-IPV |  | Unvaccinated mothers |  | TdaP<sub>5</sub>-IPV:DTaP<sub>3</sub>-IPV | TdaP<sub>3</sub>-IPV:None | TdaP<sub>5</sub>-IPV:None |
| --- | --- | --- | --- | --- | --- | --- | --- | --- | --- | --- |
|  |  | N | GMC (95% CI) | N | GMC (95% CI) | N | GMC (95% CI) | GMR (95% CI) | GMR (95% CI) | GMR (95% CI) |
| PT | Birth | 57 | 50.06 (37.55 - 66.75) | 53 | 35.84 (26.73 - 48.05) |  |  | 0.72 (0.48 - 1.07) |  |  |
|  | 2 months | 61 | 16.82 (12.64 - 22.37) | 70 | 10.7 (8.07 - 14.18) | 27 | 2.64 (1.77 - 3.94) | 0.64 (0.43 - 0.94) | 6.36 (3.82 - 10.61) | 4.05 (2.45 - 6.68) |
|  | 5 months | 62 | 30.72 (26.32 - 35.84) | 65 | 33.55 (28.89 - 38.96) | 45 | 43.2 (35.5 - 52.58) | 1.09 (0.88 - 1.35) | 0.71 (0.56 - 0.9) | 0.78 (0.61 - 0.98) |
|  | 13 months | 61 | 5.07 (3.95 - 6.51) | 63 | 6.05 (4.56 - 8.01) | 44 | 6.56 (4.79 - 9.00) | 1.19 (0.82 - 1.73) | 0.77 (0.51 - 1.16) | 0.92 (0.62 - 1.38) |
|  | 5:2 months | 60 | 1.89 (1.33 - 2.69) | 65 | 3.25 (2.29 - 4.62) | 27 | 14.1 (8.67 - 22.93) | 1.72 (1.05 - 2.81) | 0.13 (0.07 - 0.25) | 0.23 (0.12 - 0.43) |
| FHA | Birth | 57 | 245.79 (190.72 - 316.76) | 53 | 97.44 (78.44 - 121.03) |  |  | 0.4 (0.29 - 0.55) |  |  |
|  | 2 months | 61 | 66.37 (51.95 - 84.79) | 70 | 31.74 (26.05 - 38.68) | 27 | 5.97 (4.02 - 8.87) | 0.48 (0.35 - 0.65) | 11.12 (7.36 - 16.79) | 5.32 (3.55 - 7.96) |
|  | 5 months | 62 | 54.52 (47.75 - 62.25) | 65 | 50.67 (43.92 - 58.45) | 45 | 67.31 (55.02 - 82.34) | 0.93 (0.77 - 1.13) | 0.81 (0.65 - 1.01) | 0.75 (0.6 - 0.94) |
|  | 13 months | 61 | 14.01 (11.25 - 17.43) | 63 | 18.07 (13.81 - 23.64) | 44 | 22.2 (17.15 - 28.72) | 1.29 (0.92 - 1.81) | 0.63 (0.44 - 0.91) | 0.81 (0.57 - 1.17) |
|  | 5:2 months | 60 | 0.83 (0.65 - 1.05) | 65 | 1.67 (1.3 - 2.15) | 27 | 10.37 (6.31 - 17.04) | 2.02 (1.43 - 2.84) | 0.08 (0.05 - 0.13) | 0.16 (0.1 - 0.26) |
| PRN | Birth | 56 | 250.13 (160.52 - 389.77) | 52 | 230.53 (151.1 - 351.7) |  |  | 0.92 (0.51 - 1.68) |  |  |
|  | 2 months | 62 | 62.51 (39.17 - 99.75) | 70 | 83.02 (59.29 - 116.25) | 27 | 2.63 (1.44 - 4.82) | 1.33 (0.76 - 2.32) | 23.73 (11.46 - 49.15) | 31.52 (15.41 - 64.46) |
|  | 5 months | 62 | 65.88 (55.66 - 77.96) | 65 | 53.32 (42.59 - 66.76) | 44 | 73.85 (54.41 - 100.22) | 0.81 (0.61 - 1.08) | 0.89 (0.64 - 1.24) | 0.72 (0.52 - 1) |
|  | 13 months | 60 | 10.47 (8.18 - 13.42) | 63 | 7.04 (5.33 - 9.30) | 40 | 11.24 (8.4 - 15.04) | 0.67 (0.47 - 0.97) | 0.93 (0.62 - 1.39) | 0.63 (0.42 - 0.93) |
|  | 5:2 months | 61 | 1.06 (0.64 - 1.75) | 65 | 0.65 (0.45 - 0.93) | 27 | 26.54 (11.81 - 59.62) | 0.62 (0.34 - 1.12) | 0.04 (0.02 - 0.09) | 0.02 (0.01 - 0.05) |
| FIM | Birth | 56 | 34.09 (22.77 - 51.04) | 53 | 282.36 (177.32 - 449.62) |  |  | 8.28 (4.55 - 15.09) |  |  |
|  | 2 months | 61 | 9.82 (6.71 - 14.37) | 70 | 85.5 (59.59 - 122.67) | 27 | 5.2 (2.94 - 9.21) | 8.71 (5.2 - 14.58) | 1.89 (0.96 - 3.71) | 16.43 (8.47 - 31.88) |
|  | 5 months | 62 | 3.64 (2.77 - 4.79) | 65 | 18.47 (13.58 - 25.12) | 45 | 2.32 (1.73 - 3.12) | 5.07 (3.38 - 7.61) | 1.57 (1.02 - 2.41) | 7.95 (5.19 - 12.16) |
|  | 13 months | 61 | 1.54 (1.26 - 1.87) | 63 | 1.49 (1.24 - 1.79) | 44 | 1.21 (0.92 - 1.60) | 0.97 (0.75 - 1.26) | 1.27 (0.93 - 1.72) | 1.23 (0.91 - 1.67) |
|  | 5:2 months | 60 | 0.38 (0.3 - 0.48) | 65 | 0.22 (0.19 - 0.26) | 27 | 0.49 (0.33 - 0.71) | 0.58 (0.44 - 0.76) | 0.79 (0.55 - 1.14) | 0.46 (0.32 - 0.66) |

Abbreviations: DdaP<sub>3</sub>-IPV: low dose diphtheria, tetanus, acellular pertussis (three pertussis antigens), inactivated poliovirus; TdaP<sub>5</sub>-IPV: low dose diphtheria, tetanus, acellular pertussis (five pertussis antigens), inactivated poliovirus; GMC: geometric mean concentration; CI: confidence interval; GMR: geometric mean ratio; PT: pertussis toxin FHA: filamentous haemagglutinin; FIM: fimbriae types 2 and 3; PRN: pertactin

Compared to infants in the control group, infants born to vaccinated women had higher GMCs of anti-PT, FHA, FIM 2&3 and PRN IgG (GMR 4.05 [2.45-6.68], 5.32 [3.55-7.96], 16.43 [8.47-31.88], 31.52 [15.41-64.46] respectively for DTaP_5_-IPV group; GMR 6.36 [3.82-10.61], 11.12 [7.36-16.79], 1.89 [0.96-3.71], 23.73 [11.46-49.15] respectively for TdaP_3_-IPV group. [INSERT TABLE 4 – see end of manuscript for table]

### Infant antibody concentrations at 5 months of age (Table 4)

Following infant primary vaccination at 2, 3, and 4 months, infants born to women vaccinated with TdaP_5_-IPV had similar GMCs of anti-PT, FHA and PRN as infants born to women vaccinated with TdaP_3_-IPV, but higher anti-FIM 2&3 IgG concentrations.

The GMC of anti-PT IgG in infants born to TdaP_5_-IPV or TdaP_3_-IPV vaccinated mothers was lower than that in infants born to unvaccinated mothers (GMR: 0.71 [0.56-0.90] and 0.78 [0.61-0.98], respectively). For anti-FHA IgG, this effect was only seen in infants born to TdaP_5_-IPV vaccinated mothers (GMR 0.75 [0.6-0.94]).

GMCs of anti-PT IgG increased from pre- to post-primary vaccination for all infants (TdaP_5_-IPV group: fold change 1.89 [1.33-2.69]; TdaP_3_-IPV group: 3.25 [2.29-4.62]; unvaccinated group: 14.1 [8.67-22.93]). However, there was no significant difference in the fold-change in IgG to any pertussis antigens from pre-to post-primary vaccination in infants born to mothers receiving TdaP_5_-IPV compared to those infants receiving TdaP_3_-IPV.

### Infant antibody concentrations at 13 months of age (Table 4)

At 13 months of age, GMCs of anti-PT, FHA and FIM 2&3 were similar in those infants born to TdaP_3_-IPV and TdaP_5_-IPV vaccinated mothers, whereas GMCs of anti-PRN IgG was lower in infants born to TdaP_5_-IPV vaccinated mothers compared to infants born to DTaP_3_-IPV vaccinated mothers (GMR 0.67 [0.47-0.97]). Compared to infants whose mothers received neither vaccine in pregnancy, infants born to TdaP_3_-IPV vaccinated mothers had lower GMCs of anti-FHA (GMR 0.63 0.44-0.91). For anti-PRN IgG, infants born to TdaP_5_-IPV vaccinated mothers had lower concentrations compared to unvaccinated infants (GMR 0.63 [0.42-0.93]).

### Effect of GMC of pertussis-specific IgG at 2 months of age on GMC post primary vaccination and at 13 months of age

A higher GMC of anti-PT IgG at 2 months of age was associated with a lower GMC of anti-PT IgG at 5 months of age; the fold change of the post-primary GMC of anti-PT IgG was 0.92 (0.87-0.98) per two-fold change in the pre-primary GMC of anti-PT IgG. This effect was not observed at 13 months of age. A similar effect was seen for anti-FHA IgG at 13 months of age (fold change in anti-FHA IgG GMC at 13 months of age: 0.87 [95% CI 0.77-0.99] per two-fold change in pre-primary GMC), but not at 5 months of age (fold change in post-primary anti-FHA IgG GMC: 1.07 [95% CI 0.99-1.14] per 2-fold change in pre-primary GMC). Conversely, GMCs of anti-FIM 2&3 IgG was higher post-primary vaccination with lower concentrations pre-vaccination (fold change in anti-FIM IgG GMC at 5 months of age: 1.61 [95% CI 1.55 – 1.68] per two-fold change in pre-primary GMC).

### Safety

Overall, there were 5 maternal SAEs: 2 in women in the TdaP_5_-IPV group; 2 in TdaP_3_-IPV; one in an unvaccinated mother. There were 10 SAEs in infants born to women in the TdaP_5_-IPV group; 4 in infants born to women in the TdaP_3_-IPV group and 3 in infants in the control group. No SAEs were assessed as related to maternal or infant vaccination.

## Discussion

Pertussis vaccination in pregnancy is an effective and safe strategy for the prevention of pertussis in early infancy (2,4,11-27). However, blunting of the infant’s response to primary vaccination, may occur in the context of high concentrations of maternally-derived antibody. Differences in the pertussis antigen content of vaccines recommended in pregnancy might result in differential blunting of the infant’s response to primary immunization and therefore in the persistence of antibody, and of protection against pertussis, through early childhood. This is particularly important in countries, such as the UK and many low- and middle-income countries, where a pertussis-booster dose is not given until pre-school age.

To our knowledge, this is the first randomized clinical trial of different pertussis-containing vaccines in pregnancy. Other studies have examined only TdaP_5_(Adacel, Sanofi Pasteur) (4,5,7,8,28), TdaP_5_-IPV (REPEVAX, Sanofi Pasteur) (3,29) or TdaP_3_ (BOOSTRIX, GSK) (6,30,31) and found differing impacts on infant responses to primary and booster vaccinations, but no study has compared these vaccines directly. This is an important consideration for countries which recommend pertussis vaccination in pregnancy or that are considering implementing such a programme.

Passive infant immunity to pertussis is contingent on efficient transplacental passage of antibody; it is reassuring therefore that there is no difference in the PTR in women vaccinated with either vaccine. As shown in other studies, the newborn infant concentration of antibody in this study was greater than the maternal concentration, with a PTR of greater than one for all antigens (4,6-8,29,32). Multivariate analysis showed that time from third trimester vaccination to delivery was positively associated with PTR. This may be because of the effects of cumulative exposure to the maternal antibody, however placental transfer also becomes more efficient with advancing gestational age (33). The narrow time window of vaccination and the very small number of preterm deliveries made it hard to separate the influence of these factors.

At two months of age, significantly higher concentrations of pertussis-specific antibody were found in infants born to vaccinated women, compared to infants born to unvaccinated women, consistent with the high maternal vaccine efficacy observed in the UK, Australia and the United States (2,13,34). Infants of TdaP_3_-IPV vaccinated mothers had higher concentrations of anti-PT and FHA IgG compared to infants born to TdaP_5_-IPV vaccinated mothers, consistent with the increased quantity of pertussis toxoid and FHA in TdaP_3_-IPV compared to TdaP_5_-IPV. Only TdaP_5_-IPV contains FIM2&3 antigens, therefore infants born to TdaP_5_-IPV vaccinated mothers had higher concentrations of anti-FIM IgG pre-primary vaccination. There was no difference in anti-PRN IgG GMC, reflecting the similar amount of PRN antigen in both vaccines. Although there is no correlate of protection for pertussis disease, higher levels of pertussis specific antibodies are associated with protection from disease, in particular anti-PT and PRN IgG (35-38).

Whilst a higher concentration of pertussis-specific antibody is likely to correlate with clinical protection early in infancy, it might also attenuate infant vaccine responses. Reassuringly, we found no significant difference in the concentration of anti-PT, FHA or PRN IgG post-primary vaccination in infants born to TdaP_5_-IPV compared to infants born to TdaP_3_-IPV vaccinated women, suggesting that either vaccine may be employed within a national antenatal programme, with no difference in blunting seen. When compared to infants born to unvaccinated women, blunting was observed for anti-PT IgG responses, although this effect was not sustained at the end of the first year of life. A modest reduction in anti-FHA IgG was also seen in infants born to TdaP_5_-IPV mothers compared to infants born to unvaccinated mothers, but this was not observed at 13 months of age. We could not assess whether high concentrations of maternally-derived anti-FIM IgG would have blunted responses to primary infant immunization with a five component acellular pertussis (aP) vaccine, since infants in this study did not receive a FIM-containing aP vaccine. However, an earlier non-randomized study did suggest that blunting of the anti-FIM 2&3 response can also occur (3).

Whilst the blunting effect observed was statistically significant, it is important to note that the absolute difference in GMCs was small. It is particularly reassuring that UK epidemiological data did not show an excess of pertussis cases in infants born to vaccinated compared to unvaccinated women following infant primary vaccination (2). Some studies have also shown blunting post-primary vaccination (5,6,8,31); however others have not shown a significant blunting effect (4,5,28,29). In countries where a booster dose is given in the second year of life, some studies have shown persisting evidence of blunting (8,30,31), whilst others have not shown any difference post booster (4,5,28). The latter were more limited in size or had not demonstrated significant blunting post-primary vaccination. Factors which may influence blunting include maternal vaccine received, timing of infant vaccination and the pertussis antigen component of infant vaccination, both in terms of the number and amount of antigen.

### Strengths and limitations

This randomized clinical trial is the first to compare the effect of vaccination with either TdaP_5_-IPV or TdaP_3_-IPV in pregnancy on transplacental transfer of antibody and the serological response to infant primary immunization and the concentration of vaccine specific antibody at 13 months of age.

We did not include a randomized control group as this would have been unethical in the context of national recommendations for antenatal pertussis vaccination. We recruited the control group of women and infants in the postnatal period to ensure that women were not discouraged from receiving a pertussis-containing vaccine in pregnancy. We had difficulty recruiting to the control group, therefore included data from infants included in another study, carried out at the same sites using similar protocols and with samples analysed using identical methods in the same laboratory.

The study commenced towards the beginning of the antenatal pertussis programme and most women received a pertussis-containing vaccine for the first time in the current pregnancy, further studies should examine the effects of repeat doses of vaccine in subsequent pregnancies.

## Conclusion

We provide robust evidence to suggest that either TdaP_5_-IPV or TdaP_3_-IPV vaccines may be used in pregnancy, with no differential effect on the protection afforded against pertussis by infant primary immunization with an aP_3_ vaccine or that sustained into the second year of life. This data may provide reassurance that either TdaP_5_-IPV or TdaP_3_-IPV may be used within national pertussis vaccination programmes in pregnancy.

## Supporting information

Additional file 1: protocol

## Data Availability

The datasets analysed during the current study may be available from the corresponding author on reasonable request.

## Abbreviations

aP: acellular pertussis
DT: diphtheria toxoid
ELISA: enzyme-linked immunosorbent assays
FHA: filamentous haemagglutinin
FIM: fimbriae
GMC: geometric mean concentration
GMR: geometric mean ratio
GSK: GlaxoSmithKline
Hib: *Haemophilus influenza*e type b
IgG: immunoglobulin G
iMAP2: immunising Mums Against Pertussis
IPV: inactivated poliovirus
IU: International Units
LLOD: lower limit of detection
Men C: meningococcal serogroup C
PRN: pertactin
PT: pertussis toxin
PTR: placental transfer ratio
SAE: serious adverse event
TdaP_3_-IPV: low dose diphtheria toxoid, tetanus toxoid, acellular pertussis [three antigens] and inactivated polio
TdaP_5_-IPV: low dose diphtheria toxoid, tetanus toxoid, acellular pertussis [five antigens] and inactivated polio
TT: tetanus toxoid
UK: United Kingdom
WHO: World Health Organization

## Declarations

### Ethics approval and consent to participate

The study was approved by the MHRA, NHS Health Research Authority and City & East Research Ethics Committee (14/LO/0141). All participants provided informed written consent.

### Consent for publication

Not applicable

## Author contributions

Study concept and design: CEJ, PTH and EM

CEJ, NA, AC, PTH and EM had full access to all the data in the study and takes responsibility for the integrity of the data and the accuracy of the data analysis

BH, MM, AE and HC had management responsibilities over the laboratory which performed all of the testing and therefore checked and verified all the laboratory data for validity and accuracy.

Critical revision of the manuscript for important intellectual content: All authors Statistical analysis: NA

Obtained funding: EM

Administrative, technical or material support: All authors

## Acknowledgements

We thank the mothers and infants who took part in this study, the Vaccine Research Nurses in Hertfordshire and Gloucestershire for their assistance in the recruitment and follow up of the study participants and data management, the Cord Blood team at St George’s University Hospitals NHS Foundation Trust for the support in obtaining cord blood samples from participants, the scientists at Public Health England, Porton for testing the samples, Pauline Waight for her help with data management, Dr Shamez Ladhani and Dr Gayatri Amirthalingam for helpful discussions on study design and the clerical staff in the Immunization and Countermeasures Division for study administration and data entry.

## Additional materials

Additional file 1: Protocol (.pdf).

Full trial protocol

Additional file 2: Statistical analysis plan (.pdf)

Full statistical analysis plan

**Blood samples were not collected at birth from infants born to women not receiving a pertussis-containing vaccine in pregnancy as these women and infants were only recruited in the postnatal period, therefore corresponding cells are empty**

