## Additional file 1: protocol for "A phase IV, multi-centre, randomized clinical trial comparing two pertussis-containing vaccines in pregnant women in England and vaccine responses in their infants"

### (immunising Mums Against Pertussis, iMAP2)

| | |
|---|---|
| **Short Title:** | immunising Mums Against Pertussis 2 |
| **Code:** | iMAP2 |
| **PHE Protocol No:** | |
| **Eudract Number:** | 2013-004495-34 |
| **Protocol Version:** | 9 |
| **Date:** | 10 Jan 2019 |
| **Sponsor**: | Public Health England, 2nd Floor, 151 Buckingham Palace Road, London |
| **Chief Investigator:** | Professor Elizabeth Miller, Public Health England, Colindale, London |
| **Site Principal investigator:** | Professor Paul Heath, SGVI, St George's University of London |
| **Co-investigators:** | Dr Christine Jones, Dr Shamez Ladhani, Dr Gayatri Amirthalingam, Dr Jo Southern, Dr Asma Khalil |
| **Study Statistician:** | Dr Nick Andrews |

**13 REFERENCES** .......... **41**

Index to appendices - page 44

### 1 AMENDMENT HISTORY

| Amendment No. | Protocol Version No. | Date issued | Author(s) of changes | Details of Changes made |
|---|---|---|---|---|
| *1* | *3* | *14.8.14* | *Jo Southern* | *Correction of column headings in treatment table(section 6.2)*<br>*Amendment of first infant blood sample, to be collected from 0-7 days* |
| *2* | *4* | | *Jo Southern/ Kevin Brown* | *Addition of polio testing in main protocol and appendices, as well as actions for extra vaccine doses on low antibody results* |
| *4* | *6* | *08/07/2015* | *Chrissie Jones / Jo Southern* | *Addition of Bexsero (Meningococcal group B vaccine) to vaccine schedule as per UK national immunisation schedule. Inclusion of possible testing of Men B responses if sufficient sera and funding* |

| | | | | |
|---|---|---|---|---|
| *5* | *7* | *11.1.16* | *Jo Southern* | *Inclusion of option for home visits for vaccination as well as sample collection on p.28* |
| *7* | *8* | *4.10.18* | | *Version withdrawn due to missing information with polio secondary objective – corrected to version 9 and resubmitted* |
| *8* | *9* | *10.1.19* | *Elizabeth Miller* | *Relegation of polio testing from secondary objective to exploratory analysis*<br>*Change of polio testing lab from PHE to NIBSC* |

### 2 SYNOPSIS

| | |
|---|---|
| **Study Title** | A randomised controlled trial comparing two pertussis-containing vaccines in pregnancy and vaccine responses of UK mothers and their infants |
| **Short title** | immunising Mums Against Pertussis 2 |
| **Study code** | iMAP2 |
| **Study Design** | Multi-centre randomised trial with a non-randomised control group. |
| **Study Participants** | Women in the 2nd and 3rd trimester of pregnancy and post-partum women and their infants |
| **Number of participants** | Total of 180 mother/infant pairs (130 mother/infant pairs with mother randomised to either REPEVAX or BOOSTRIX-IPV and 50 non-randomised unvaccinated women and their infants) |
| **Recruitment period** | 12 months (anticipated) |
| **Study Period** | January 2014 – January 2017 |
| **Primary Objective** | • To compare anti-pertussis toxin (PT) IgG responses following primary immunisation with an acellular pertussis- containing vaccine in infants born to mothers who received REPEVAX in pregnancy compared to infants whose mothers received BOOSTRIX-IPV in pregnancy. |
| **Secondary Objectives** | • To compare antibody responses to pertussis antigens (concentration of IgG antibody to PT, pertactin (PRN), filamentous haemagglutinnin (FHA) and fimbrial antigens 2 and 3 (FIM 2 and 3]), tetanus toxoid, diphtheria toxoid at birth amongst infants born to mothers who received REPEVAX in pregnancy compared to infants whose mothers received BOOSTRIX-IPV in pregnancy<br>• To compare antibody responses to pertussis antigens [IgG to PT, PRN, FHA and FIM 2 and 3], Hib antigen [PRP], tetanus toxoid, diphtheria toxoid; meningococcal serogroup C serum bactericidal antibody titres |

| | |
|---|---|
| | and meningococcal serogroup C-specific IgG concentrations; 13 serotype-specific pneumococcal IgG concentrations and functional pneumococcal antibody studies at 2, 5 and 13 months of age (just before and one month after primary immunization and one month after booster vaccines) in infants born to mothers who received REPEVAX in pregnancy compared to infants whose mothers received BOOSTRIX-IPV in pregnancy and compared to infants whose mothers did not receive pertussis vaccination in pregnancy<br>• To determine concentrations of antibodies to pertussis antigens [IgG to PT, PRN, FHA and FIM 2 and 3] and tetanus toxoid, diphtheria toxoid, in pregnant women prior to administration of the REPEVAX or BOOSTRIX-IPV vaccine and at the time of delivery<br>• To estimate the placental transfer ratio of antibodies to pertussis antigens [IgG to PT, PRN, FHA and FIM 2 and 3] and tetanus toxoid, diphtheria toxoid, in mothers who received REPEVAX in pregnancy compared to mothers who received BOOSTRIX-IPV in pregnancy<br>• To determine persistence of pertussis IgG (PT, PRN, FHA and FIM 2 and 3) in mothers at 13 months after delivery in mothers who received either REPEVAX, BOOSTRIX-IPV or no pertussis-containing vaccine |
| **Exploratory analyses** | To investigate whether there is an effect of maternal immunisation with a TdaP/IPV vaccine on infants' responses to inactivated polio vaccine |
| **Inclusion Criteria** | Pregnant women who, at the time of enrolment<br>• are aged 16- 45 years |
| **Exclusion Criteria** | Participant may not be included in the study if <u>any</u> of the following apply:<br>All women:<br>• Bleeding disorder<br>• Receipt of any pertussis containing vaccine in the previous 12 months<br>Women to be vaccinated only (i.e. not the control group):<br>• Received immunoglobulin or other blood product within the preceding 3 months |

| | |
|---|---|
| | • Fulfil any of the contraindications to vaccination specified in The Green Book on Immunisation (https://www.gov.uk/government/organisations/public-health-england/series/immunisation-against-infectious-disease-the-green-book), including:<br>⋆ A confirmed anaphylactic reaction to a previous dose of diphtheria, tetanus, pertussis or poliomyelitis containing vaccine<br>⋆ A confirmed anaphylactic reaction to any component of the vaccine<br>⋆ A confirmed anaphylactic reaction to a previous dose of diphtheria, tetanus, pertussis or poliomyelitis containing vaccine |
| **Inclusion/ Exclusion criteria for infants** | Infants will be vaccinated under the routine national immunisation schedule in accordance with the criteria set out in the Department of Health "Green Book" |
| **Temporary exclusion criteria** | If the pregnant woman or the baby has an axillary/aural temperature ≥ 38°C, then vaccination and blood sampling will be postponed until resolution of fever. If the pregnant woman or baby is acutely unwell, vaccination will postponed until resolution. Blood sampling will also be postponed for seven days after completion of any antibiotic course. |
| **Intervention** | Randomisation of pregnant women to receive either REPEVAX or BOOSTRIX-IPV at 28 - 32 weeks gestation. Vaccination of infants under the routine national schedule at 2, 3,4 and 12 months of age. |
| **Primary Endpoint** | • Fold-difference in anti-PT IgG GMC in infants at 5 months of age whose mothers received either REPEVAX or BOOSTRIX-IPV |
| **Secondary Endpoints** | • Geometric mean concentration (GMC) of IgG to pertussis antigens (PT, PRN, FHA and FIM 2 and 3), tetanus toxoid and diphtheria toxoid in infants at birth whose mothers received either REPEVAX or BOOSTRIX-IPV |

| | |
|---|---|
| | • GMC of IgG to pertussis antigens (PT, PRN, FHA and FIM 2 and 3); anti-PRP IgG [Hib antigen] GMC and proportion of infants with concentration ≥ 0.15 and ≥ 1.0 ug/ml; anti-tetanus toxoid IgG GMC; anti-diphtheria toxoid IgG GMC; meningococcal serogroup C serum bactericidal antibody titres (GMT) and proportion of infants with titres ≥ 8 and ≥128; meningococcal serogroup C-specific IgG GMC; 13 serotype-specific pneumococcal IgG GMCs and functional pneumococcal antibody studies in infants at 2, 5 and 13 months whose mothers received either REPEVAX, BOOSTRIX-IPV or no pertussis-containing vaccine.<br>• GMC of IgG to pertussis antigens (PT, PRN, FHA and FIM 2 and 3), tetanus toxoid, diphtheria toxoid in mothers pre-vaccination and at delivery who received REPEVAX or BOOSTRIX-IPV.<br>• Placental transfer ratio of IgG to pertussis antigens (PT, PRN, FHA and FIM 2 and 3), tetanus toxoid, diphtheria toxoid from mothers to their infants in mothers who received REPEVAX or BOOSTRIX-IPV.<br>• Persistence of pertussis IgG (PT, PRN, FHA and FIM 2 and 3) at 13 months after delivery in mothers who received either REPEVAX, BOOSTRIX-IPV or no pertussis-containing vaccine measured as the geometric mean ratio of concentrations at 13months v delivery. |
| **Exploratory endpoints** | Comparisons in serotype-specific polio antibody levels (GMTs and percentages with titres ≥1 in 8 by microneutralization) will be made between unvaccinated mothers and their infants and vaccinated mothers and their infants with data for the latter combined across the two TdaP/IPV vaccines used for maternal immunisation |
| **Definition of end of study** | Date of reporting of the final blood test result. |

### 3 LIST OF ABBREVIATIONS

| | |
|---|---|
| AE | Adverse Event |
| CI | Chief Investigator |
| CRF | Case Report Form |
| $CRM_{197}$ | *Corynebacterium diphtheriae* $CRM_{197}$ protein |
| CT | Clinical Trial |
| CTA | Clinical Trial Authorisation |
| ELISA | Enzyme-linked Immunosorbent Assay |
| EMA | European Medicine Agency |
| FDA | Food Drug Administration |
| FHA | Filamentous hemagglutinin |
| FIM 2/3 | Fimbriae types 2 and 3 |
| GCP | Good Clinical Practice |
| GMC | Geometric Mean Concentration |
| GMT | Geometric Mean Titre |
| GP | General Practitioner |
| GSK | GlaxoSmithKline |
| Hib | *Haemophilus influenzae* type b |
| ICH | International Conference on Harmonisation |
| IM | Intramuscular |
| IPV | Inactivated polio vaccine |
| IRAS | Integrated Research Application System |
| MCC | Meningococcal serogroup C conjugate |
| Men B | *Neisseria meningitidis* serogroup B |
| MenC | *Neisseria meningitidis* serogroup C |

| | |
|---|---|
| µg | Micrograms |
| MHRA | Medicines and Healthcare Products Regulatory Authority |
| NHS | National Health Service |
| NRES | National Research Ethics Service |
| PCV13 | 13-valent Pneumococcal Conjugate Vaccine (Prevenar13®) |
| PHE | Public Health England |
| PI | Principal Investigator |
| PIL | Participant/Parent Information Leaflet |
| PRN | Pertactin |
| PT | Pertussis Toxin |
| SAE | Serious Adverse Event |
| SBA | Serum Bactericidal Antibody |
| SGH | St. George’s Hospital, London |
| SGUL | St. George’s, University of London |
| SGVI | St. George’s Vaccine Institute |
| SOP | Standard Operating Procedure |
| SPC | Summary of Product Characteristics |
| SUSAR | Suspected Unexpected Serious Adverse Reaction |

### 4 BACKGROUND AND RATIONALE

Childhood immunisation against pertussis has been routine in England and Wales since 1957 and has led to a dramatic decline in pertussis. Despite maintenance of high vaccine coverage of a 5-component pertussis vaccine (Pediacel®; DTaP5-Hib-IPV) pertussis continues to display 3-4 yearly cyclical peaks in activity. In 2011, an increase in laboratory confirmed cases of pertussis was observed, in excess of previous peak years. Cases of pertussis continued to increase into 2012 and Public Health England (formerly the Health Protection Agency) declared a level 3 incident in April 2012 (Health Protection News, 2012).

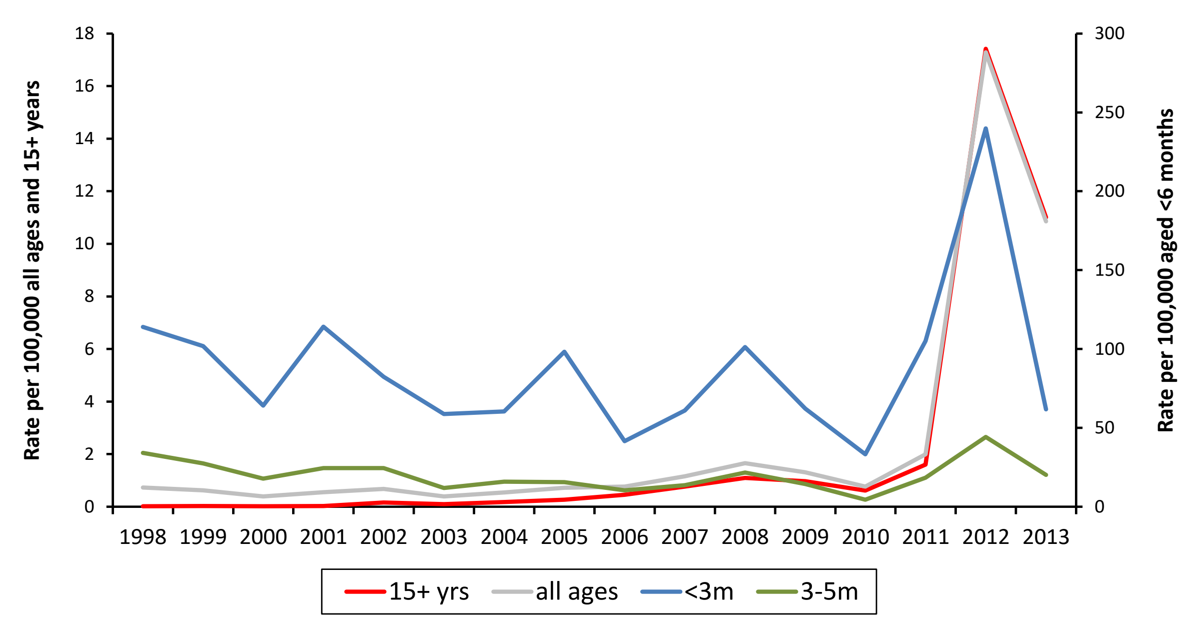

Figure 1. Incidence of laboratory-confirmed pertussis, by total case-patients aged <3 months, 3-5 months, ≥15 years, and all ages in England and Wales since 1998. Data for 2013 only include the first five months (January-May).

In 2012, there were 9,711 laboratory confirmed cases in England and Wales, almost nine-fold higher than 2011 with confirmed cases peaking at 1,628 in October (Health Protection Reports, 2013). Young infants were disproportionately affected, with 429 confirmed cases in <3 month-olds (239.8/100,000) peaking at 55 confirmed cases in August. During 2012, 14 infant deaths were reported, the highest number of annual infant deaths in more than 15 years. As a result, the Department of Health announced a temporary programme to offer pertussis vaccination to all pregnant women in the UK during the third trimester from September 2012 in order to protect young infants (Department of Health, Health, 2012).

Acceptance of the temporary pertussis vaccination programme in pregnancy has been high; coverage peaked at 78% in January 2013 and has been maintained at 68% since this time (Unpublished data, Amirthalingam, 2013). There is evidence that the maternal vaccination programme has been effective in preventing infant disease (Unpublished data, Amirthalingam, 2013). Laboratory confirmed cases of pertussis have declined in all age groups less than 20 years of age since November 2012, in line with seasonal trends, however the decrease amongst the youngest infants has been more pronounced than in the older age groups. Amongst infants less than 2 months of age, a 70-73% decrease has been observed between January and June 2012 and the same period in 2013, compared to a decrease of 29% in infants 3-11 months and 15% in 5-19 year olds. This decrease in pertussis notifications is supported by a decrease in hospital admissions and deaths in young infants. In 2013 only one death has occurred in an unvaccinated mother compared to 14 infant deaths in 2012. Vaccine efficacy is estimated at 92% (95% CI: 85%-96%). This data has contributed to the JCVI decision to continue with the maternal vaccination programme into 2014 (Department of Health, 2013).

Maternal pertussis vaccination is likely to protect the infant from pertussis in two ways: transplacental transfer of antibody and prevention of pertussis in the mother around the time of delivery and therefore avoidance of pertussis exposure in the infant.

Antibody is actively transferred across the placenta from mother to child. A number of studies have provided evidence supporting the existence of efficient transplacental transfer of pertussis antibodies (Gall et al., 2011; Leuridan et al., 2011). Cord blood from newborn infants whose mothers received TdaP during or before pregnancy had higher concentrations of pertussis antibodies when compared with cord blood from newborn infants of unvaccinated mothers (Halperin et al., 2011).

However, evidence from some studies suggests that maternal pertussis antibodies can inhibit pertussis-specific antibody responses of infants receiving primary immunisations with DTaP, an effect known as “blunting” (van Savage,1990; England, 1995). As there are no specific correlates of protection for pertussis (i.e. the level of pertussis antibody considered to confer protection against infection), the clinical relevance of blunting is uncertain (ACIP, 2011). It is expected that any blunting that occurs is short-lived because maternal antibodies decline rapidly in the infant and, although the antibody response to the first dose of a pertussis-containing vaccine may be lower in infants whose mothers received a pertussis vaccine in pregnancy, an adequate immune response should be achieved in all infants after completion of the primary immunisation schedule (de Greef, 2010).

Currently, two studies are underway in the US and Canada to explore the impact of maternal vaccination with TdaP in the third trimester of pregnancy on the immune response to infants receiving DTaP at 2, 4 and 6 months (ClinicalTrials.gov Identifier: NCT00707148; ClinicalTrials.gov Identifier: NCT00553228). Recent analysis of interim data from one trial found elevated anti-pertussis antibody concentrations at birth and before the first dose of vaccine in infants whose mothers received TdaP compared to those whose mothers received Td (Gall et al., 2011). After the third dose of vaccine the former group had lower pertussis antibody concentrations. Most children in this group were considered to have acceptable antibody responses when the results were reviewed by the US Advisory Committee on Immunization Practices (ACIP), which concluded that the short duration of blunting and potential risk of shifting the burden to older infants was outweighed by the potential protection afforded by higher maternal antibodies in early infancy (Gall et al., 2011). Whether these results are applicable to the UK is not known, since the UK has introduced a dTap-IPV vaccine (REPEVAX, Sanofi-Pasteur MSD) in pregnancy and infants are immunised at an accelerated 2-3-4 month schedule with a different pertussis-containing combination vaccine.

In addition to blunting, it is possible that high levels of maternally-derived antibody in the infant at birth may cause a reduction in the infant's response to other vaccine antigens as a result of carrier-mediated suppression of antibody responses. A single study carried out in The Gambia showed that pregnant women who were immunized with tetanus toxoid vaccine 1-5 weeks before being vaccinated with a polysaccharide-tetanus protein conjugate vaccine had lower anti-PRP responses than women who had not previously received tetanus vaccination in the same pregnancy (Mulholland, 1996). Cord blood levels of anti-PRP were also reduced when the mother had received prior tetanus vaccination. They observed a negative correlation between pre-existing maternal anti-tetanus antibody concentrations and subsequent cord anti-PRP responses. REPEVAX and BOOSTRIX-IPV both contain tetanus toxoid and therefore it is conceivable that vaccination with one of these vaccines during pregnancy would lead to increased anti-tetanus antibody in the infant which may affect the infants response to PRP covalently bound to tetanus toxoid (as found in the infant vaccine Infanrix-IPV/Hib). It is therefore important to assess infant responses to other vaccine antigens.

A further rationale for monitoring responses to other antigens in the primary infant vaccine schedule comes from studies of neonatal vaccination with pertussis containing vaccines. Knuf et al observed that whilst vaccination with a neonatal dose of acellular pertussis induced high concentrations of antibody to pertussis antigens, the GMC of anti-PRP was lower following primary infant vaccinations (Knuf, 2008). Furthermore, Halasa et al found that a neonatal dose of DTaP was associated with reduced responses to pertussis antigens and diphtheria toxoid compared to controls (Halasa 2008). Whilst the cellular response to neonatal vaccination is not analogous to that of the passive immunity derived from maternal vaccination these studies urge a broad assessment of how high levels of antibody at birth might affect infant responses to both related antigens and well as unrelated antigens.

REPEVAX is the currently available vaccine for maternal pertussis vaccination in the UK; however, BOOSTRIX-IPV (GSK) is licensed for adolescent boosting as it is of proven efficacy in this age group and will be used in the national campaign from the spring of 2014 as a result of a tendering process that was unable to take account of any differences between the vaccines in terms of passive immunity conferred to the infant or potential blunting of infants' responses to primary immunisation. There are no studies directly comparing REPEVAX and BOOSTRIX-IPV in pregnancy. Both vaccines contain the same quantity of diphtheria toxoid, tetanus toxoid and inactivated polio, but there are differences in the pertussis antigens. REPEVAX contains 2.5 micrograms (μg) of pertussis toxoid (PT), 5μg of filamentous haemagglutinin (FHA), 3μg of pertactin (PRN) and 5μg of fimbriae (FIM) types 2 and 3. BOOSTRIX-IPV contains 8μg of PT, 8μg of FHA and 2.5μg of PRN, but does not contain the FIM antigens. These differences may lead to different immunogenicity of the vaccines in pregnant women as well as different antibody concentrations in newborns at birth, before and after primary immunisation. It is therefore important to assess these two vaccines in pregnancy to identify any differences in the response to vaccination in infants. This is essential to assess any effect of pertussis-containing vaccines on infants' primary immunisations. This study therefore aims to assess immunological responses following REPEVAX and BOOSTRIX-IPV in pregnant women and their infants and compare them to a control group of women who declined to receive a pertussis-containing vaccine during pregnancy.

### 5 OBJECTIVES

#### 5.1 Primary Objective

To compare anti-pertussis toxin (PT) IgG responses following primary immunisation with an acellular pertussis- containing vaccine in infants born to mothers who received REPEVAX in pregnancy compared to infants whose mothers received BOOSTRIX-IPV in pregnancy.

#### 5.2 Secondary Objectives

**Infant**

- To compare antibody responses to pertussis antigens (concentration of IgG antibody to PT, pertactin (PRN), filamentous haemagglutinnin (FHA) and fimbrial antigens 2 and 3 (FIM 2 and 3]), tetanus toxoid and diphtheria toxoid at birth amongst infants born to mothers who received REPEVAX in pregnancy compared to infants whose mothers received BOOSTRIX-IPV in pregnancy

  To compare antibody responses to pertussis antigens [IgG to PT, PRN, FHA and FIM 2 and 3], Hib antigen [PRP], tetanus toxoid and diphtheria toxoid; meningococcal serogroup C serum bactericidal antibody titres and meningococcal serogroup C-specific IgG concentrations; 13 serotype-specific pneumococcal IgG concentrations and functional pneumococcal antibody studies at 2, 5 and 13 months of age (just before and one month after primary immunization and one month after booster vaccines) in infants born to mothers who received REPEVAX in pregnancy compared to infants whose mothers received BOOSTRIX-IPV in pregnancy and compared to infants whose mothers who did not receive pertussis vaccination in pregnancy

**Mother**

- To determine concentrations of antibodies to pertussis antigens [IgG to PT, PRN, FHA and FIM 2 and 3] and tetanus toxoid and diphtheria toxoid in pregnant women prior to administration of the REPEVAX or BOOSTRIX-IPV vaccine and at the time of delivery
- To estimate the placental transfer ratio of antibodies to pertussis antigens [IgG to PT, PRN, FHA and FIM 2 and 3] and tetanus toxoid and diphtheria toxoid, in mothers who received REPEVAX in pregnancy compared to mothers who received BOOSTRIX-IPV in pregnancy

- To compare concentrations of antibodies to pertussis antigens [IgG to PT, PRN, FHA and FIM 2 and 3] at 12-13 months post-delivery in women who received REPEVAX or BOOSTRIX-IPV vaccine and to those women who did not receive pertussis vaccination in pregnancy

##### 5.3 Exploratory analyses

- To investigate whether there is an effect of maternal immunisation with a TdaP/IPV vaccine on infants' responses to inactivated polio vaccine

#### 6 STUDY DESIGN

##### 6.1 Study design

This clinical trial will prospectively recruit approximately 130 pregnant women attending their routine antenatal appointments at St. George's Healthcare NHS trust in South West London or their GP surgery in Hertfordshire, Gloucestershire or South London. Of the 130 (+/-10) women recruited in the antenatal period, 65 (+/-10) women will be randomised to receive REPEVAX and 65 (+/-10) will be randomised to receive BOOSTRIX-IPV. 50(+/-10) women who declined to receive pertussis vaccination in pregnancy will also be recruited as a control group. These women will be recruited in the post-natal period using a separate information pack offering to test their baby's vaccine responses and providing booster doses of meningococcal C, Hib or pneumococcal vaccine(s) if responses are sub-optimal*. Written informed consent will be obtained prior to enrolment. The women opting to receive pertussis vaccination in pregnancy will be vaccinated by the study team in accordance with the inclusion/ exclusion criteria for the trial and the schedule set out in the table below. The study team will follow up the mothers and infants and administer all the routine infant vaccinations during the study period. The sampling schedule is set out in section 6.2.

*An extra dose of polio vaccine may be offered if the child is known to be travelling to a polio endemic country prior to the pre-school booster, this will be given in the form of Revaxis (dT-IPV). Otherwise, all children will routinely be offered a polio containing vaccine at the pre-school booster as per the national immunisation schedule.

#### 6.2 Study Schedule

| **Visit** | **V1 Pregnancy** | **V2 Birth** | **V3 2 months** | **V4 3 months** | **V5 4 months** | **V6 5 months** | **V7 12 months** | **V8 13 months** |
|---|---|---|---|---|---|---|---|---|
| **Visit Window** | 28-32 weeks | 0-7 days hours after birth | 49-84 days of age | 21-42 days after visit 3 | 21-42 days after visit 4 | 21-42 days after visit5 | 353-390 days of age | 21-42 days after visit 7 |
| **Vaccinated mothers: Maternal vaccination** | REPEVAX Or BOOSTRIX-IPV | | | | | | | |
| **Vaccinated mothers: Maternal Blood Test** | ✓ **Blood MA** | ✓ **Blood MB** | | | | | | ✓ **Blood MC** |
| **Unvaccinated mothers** | | | | | | | | ✓ **Blood MC** |
| **Infants born to vaccinated mothers: vaccination** | | | Infanrix-IPV+Hib Prevenar13 Bexsero Rotarix | Infanrix-IPV+Hib NeisVac-C Rotarix | Infanrix-IPV+Hib Prevenar13 Bexsero | | Menitorix Prevenar13 Bexsero | MMR** |
| **Infants born to vaccinated mothers: Baby Blood Test** | | ✓ **Blood BA* (Cord blood or venepuncture)** | ✓ **Blood BB** | | | ✓ **Blood BC** | | ✓ **Blood BD** |
| **Infants born to unvaccinated mothers: Vaccination** | | | Infanrix-IPV+Hib Prevenar13 Bexsero Rotarix | Infanrix-IPV+Hib NeisVac-C Rotarix | Infanrix-IPV+Hib Prevenar13 Bexsero | | Menitorix Prevenar13 Bexsero | MMR** |
| **Infants born to unvaccinated mothers: Baby blood test** | | | ✓ **Blood BB** | | | ✓ **Blood BC** | | ✓ **Blood BD** |

Table2. Study Schedule.

*Cord blood will be collected where possible, otherwise peripheral blood sampling

** Infants may receive one of 2 licensed MMR vaccines depending on local availability: Priorix or MMR®II. The MMR can be given at the 12 month visit alongside other vaccines, as per the national schedule, if the family wishes.

#### 6.3 Study Participants

Pregnant women opting to enrol in the study and their babies must fulfil all inclusion criteria and none of the exclusion criteria in order to be eligible to participate:

##### 6.3.1 Inclusion Criteria for pregnant women (vaccinated group)

Pregnant women at any stage of their pregnancy who, at the time of enrolment

- are aged 16- 45 years

##### 6.3.2 Exclusion criteria for pregnant women (vaccinated group)

Participant may not be included in the study if any of the following apply:

- Bleeding disorder
- Received immunoglobulin or other blood product within the preceding 3 months
- Fulfil any of the contraindications to vaccination specified in The Green Book on Immunisation (https://www.gov.uk/government/organisations/public-health-england/series/immunisation-against-infectious-disease-the-green-book), including:
    - A confirmed anaphylactic reaction to a previous dose of diphtheria, tetanus, pertussis or poliomyelitis containing vaccine
    - A confirmed anaphylactic reaction to any component of the vaccine
    - A confirmed anaphylactic reaction to a previous dose of diphtheria, tetanus, pertussis or poliomyelitis containing vaccine
- Receipt of any pertussis containing vaccine in the previous 12 months

##### 6.3.3 Exclusion criteria for women who declined pertussis vaccination (unvaccinated group)

- Bleeding disorder
- Receipt of any pertussis containing vaccine in the previous 12 months

##### 6.3.4 Inclusion/ Exclusion criteria for infants

Infants will be vaccinated under the routine national immunisation schedule in accordance with the criteria set out in the Department of Health "Green Book"

##### 6.3.5 Temporary Exclusion Criteria

If the pregnant woman or the baby has an axillary/oral/ aural temperature ≥ 38°C, then vaccination and blood sampling will be postponed until resolution of fever. If the pregnant woman or baby is acutely unwell, vaccination will postponed until resolution. Blood sampling will also be postponed for seven days after completion of any antibiotic course.

#### 6.4 Study procedures

##### 6.4.1 Recruitment

**Recruitment in South London**

Women attending antenatal scans or clinic appointments at St. George's Hospital will be offered a covering letter (CovLetSGH1, Appendix 1) along with a Pre-Information Leaflet containing information about the study (PreInfo1, Appendix 1). Women who contact the St. George's study team by text/phone/email/reply slip (thus indicating their interest in receiving more information about the study) will be contacted by one of the St. George's Vaccine Institute (SGVI) study staff, who will explain the purpose of the study, answer any questions relating to the study and provide written information about the study (Participant Information Leaflet, PIL, Appendix 1).

GPs in South London will also be approached to agree to the participation of their surgery in the study and will be asked to document their agreement to the study being conducted in their surgery by the completion of a protocol approval form (Appendix 1). No participants will be recruited from a surgery until this has been obtained. A site file will be provided to each participating surgery, with documents as listed in the index (Appendix). Study investigators and co-ordinators will be available to contact should health professionals or parents require any further information about the study.

GP surgeries will identify eligible pregnant women and provide them with a covering letter (CovLetSGH-GP1, Appendix 1) on behalf of the study team along with a Pre-Information Leaflet containing information about the study (PreInfo1, Appendix 1). GP surgeries may also post the information to pregnant women registered at their surgery. Women who subsequently contact the study will be contacted by one of the SGVI study team, who will explain the purpose of the study, answer any questions relating to the study and provide with written information about the study.

The control group of women who have declined to accept the currently recommended pertussis-containing vaccine in pregnancy will be recruited in the post-natal period. Women may be approached on the post-natal wards or in post-natal clinics or sent a letter by the GP (CovLetSGH-GP2 and PreInfo2, Appendix 1). They will receive a covering letter (CovLetSGH2, Appendix 1) of behalf of the study team along with a Pre-Information Leaflet (PreInfo2, Appendix 1) containing information about the study. Women indicating their interest in the study will have the opportunity to have any questions relating to the study answered and will received written information about the study (including the Participant Information Leaflet, PIL, Appendix 1).

**Recruitment in Hertfordshire and Gloucestershire**

GPs in Hertfordshire and Gloucestershire will be approached to agree to the participation of their surgery in the study and will be asked to document their agreement to the study being conducted in their surgery by the completion of a protocol approval form (Appendix 0). No participants will be recruited from a surgery until this has been obtained. A site file will be provided to each participating surgery, with documents as listed in the index (Appendix). Study investigators and co-ordinators will be available to contact should health professionals or parents require any further information.

GP surgeries will identify eligible pregnant women and provide them with a covering letter on behalf of the study team along with a Pre-Information Leaflet containing information about the study (CovLetPHE1, PreInfo 1, Appendix 1). GP surgeries may also post the information to pregnant women registered at their surgery. GP surgeries will identify women in the post-natal period who did not receive any pertussis vaccine during pregnancy and provide them with information about the study for potential recruitment into the control group study (CovLetPHE2, PreInfo2, Appendix 1). Women indicating their interest in the study will have the opportunity to have any questions relating to the study answered and will received written information about the study (including the Participant Information Leaflet, PIL, Appendix 1).

##### 6.4.2 Informed Consent

In order to take part in the study, participating mothers must sign and date the informed consent form (Appendix 1) before any study-specific procedures are performed. The participant information leaflet clearly states that the mother is free to withdraw herself and her child from the study at any time for any reason without prejudice to future care, and with no obligation to give any reason for withdrawal. All study staff are trained and experienced in obtaining informed consent for clinical trials. A copy of the signed informed consent will be given to the participant and a copy will be retained at the study site. GPs will be informed in writing of the participation of each mother and infant in the study (Appendix 0). The original signed consent form will be returned to the Chief Investigator at PHE Colindale to be stored in the participant's study file.

##### 6.4.3 Randomisation procedure

A computerised block randomisation list will be produced by the Statistician at PHE. Each centre will be allocated blocks of sequential numbers. On recruitment to the study, each subject will be allocated, in order of inclusion, the next available subject number. The study number will define the group to which the mother is assigned and which pertussis-containing vaccine she will receive.

Mothers and their infants who are recruited in the postnatal period as part of the control group will not be randomised.

##### 6.4.4 Blinding

Laboratory staff that test maternal and infant blood samples for vaccine responses will be blinded to the group allocation. Pregnant women and other study personnel will not be blinded. Though written informed consent will be taken at the first visit, continued consent to participate will be ascertained at each subsequent visit.

##### 6.4.5 Study visits

**Visit 1 (maternal vaccination)**

Study visit only includes mothers opting to receive a pertussis-containing vaccine in pregnancy.

- Provide any further information to the family as requested
- Verify inclusion and exclusion criteria with the mother (if any notes need to be checked this would be done after consent)
- Obtain written informed consent from the woman
- If the woman meets all the inclusion criteria and none of the exclusion criteria, enrol in the study and allocate the next sequential unique participant study number.
- Obtain and record medical history and concomitant medications.
- Measure axillary/oral/ aural temperature and record on source document.
    - Take blood sample (Blood MA)
    - Administer REPEVAX or BOOSTRIX-IPV according to allocation in the left upper arm
    - Observe for 20 minutes
    - Record vaccination details in research documentation in the woman's medical records at the GP surgery (Hertfordshire and Gloucestershire) or on letter to GP (St George's).
    - Instruct participant to notify study team of any serious adverse events/reactions.

- Instruct participant to notify study team as soon as the baby is born
- Provide participant with Cord Blood and Maternal Blood Collection card

**Visit 2 (within 7 days of birth)**

Study visit only includes mothers opting to receive a pertussis-containing vaccine in pregnancy and will be either a blood sample taken for the study around the time of delivery and/or a cord blood sample and may be performed whilst in hospital or at home if already discharged.

- Obtain interim history, including any serious adverse events, serious reactions, and visits to physicians or hospitalisations.
- Take blood sample from mother (Blood MB), if not taken in hospital around the time of delivery
- Obtain blood sample from baby (Blood BA), if a cord blood sample was not collected in hospital.
- Schedule Visit 3

**Visit 3 (2 months postnatal age)**

For the control group of women who did not receive a pertussis-containing vaccine in pregnancy:

- Provide any further explanation necessary about the study
- Verify inclusion and exclusion criteria
- Obtain written informed consent with the mother (if any notes need to be checked this would be done after consent)

All women and their infants:

- Obtain interim history for baby, including visits to physicians or hospitalisations and concomitant medications (if baby on antibiotics, then delay blood sampling and vaccination until at least 7 days after completion of antibiotic course, if possible)
- Take baby's axillary/aural temperature and record on source document
- Take blood sample from baby (Blood BB)
- Ensure baby fulfils criteria for vaccination according to the Green Book on Immunisation
- Vaccinate baby:
    - Infanrix-IPV+Hib into LEFT antero-lateral thigh
    - Bexsero into LEFT antero-lateral thigh
    - Prevenar 13 in RIGHT antero-lateral thigh
    - Rotarix orally

  - Paracetamol to be provided. Three doses to be given at 4-6 hourly intervals with the 1st dose given as soon as possible after Bexsero vaccination.
  - Schedule Visit 4

**Visit 4 (3 months postnatal age)**

Study visit includes all women and their infants.

- Obtain interim history for baby, including visits to physicians or hospitalisations and concomitant medications
- Take baby's axillary/aural temperature and record on source document
- Ensure baby fulfils criteria for vaccination according to the Green Book on Immunisation
- Vaccinate baby:
  - Infanrix-IPV+Hib into LEFT antero-lateral thigh
  - NeisVac-C vaccine into RIGHT antero-lateral thigh
  - Rotarix orally
- Schedule Visit 5

**Visit 5 (4 months postnatal age)**

Study visit includes all women and their infants.

- Obtain interim history for baby, including visits to physicians or hospitalisations and concomitant medications.
- Take baby's axillary/aural temperature and record on source document
- Ensure baby fulfils criteria for vaccination according to the Green Book on Immunisation
- Vaccinate baby:
  - Infanrix-IPV+Hib into LEFT antero-lateral thigh
  - Bexsero into LEFT antero-lateral thigh
  - Prevenar13 into RIGHT antero-lateral thigh
  - Paracetamol to be provided. Three doses to be given at 4-6 hourly intervals with the 1st dose given as soon as possible after Bexsero vaccination.
- Schedule Visit 6

**Visit 6 (5 months postnatal age)**

Study visit includes all women and their infants.

- Obtain interim history for baby, including visits to physicians or hospitalisations and concomitant medications (if baby on antibiotics, then delay blood sampling until at least 7 days after completion of antibiotic course, if possible)
- Take blood sample from baby (Blood BC)
- Schedule Visit 7

**Visit 7 (12 months postnatal age)**

Study visit includes all women and their infants

- Obtain interim history for baby, including visits to physicians or hospitalisations and concomitant medications
- Take baby's axillary/aural temperature and record on source document
- Ensure baby fulfils criteria for vaccination according to the Green Book on Immunisation
- Vaccinate baby:
    - Prevenar13 in RIGHT antero-lateral thigh
    - Menitorix into LEFT antero-lateral thigh
    - Bexsero into LEFT antero-lateral thigh
    - Paracetamol to be provided. Three doses to be given at 4-6 hourly intervals with the 1$^{st}$ dose given as soon as possible after Bexsero vaccination.
    - May also receive MMR at this visit if family wishes into RIGHT antero-lateral thigh

**Visit 8 (13 months postnatal age)**

Study visit includes all women and their infants

- Obtain interim history for baby, including visits to physicians or hospitalisations and concomitant medications (if baby on antibiotics, then delay blood sampling and vaccination until at least 7 days after completion of antibiotic course, if possible)
- Take baby's axillary/aural temperature and record on source document
- Take blood sample from mother (Blood MC)
- Take blood sample from baby (Blood BD)
- Vaccinate baby:
    - MMR vaccine into RIGHT antero-lateral thigh (this may have been given at 12 months )

**Other visits**

Where protective levels are not reached in the infant for polio serotypes 1, 2 or 3 in the blood samples taken at 5 months of age, or for Hib, PCV or MenC from the blood sample taken at 13 months of age, parents will be contacted to discuss the results and to recommend further dose(s) of vaccine, which will be administered by the study team. The levels to be used in this study to indicate an inadequate response are in accordance with established correlates of protection [Andrews et al., 2003; Käyhty et al., 1983] and, therefore, the recommendation of extra doses will be if:

- Polio if any of the three serotypes <1:8 by microneutralisation AND the child is known to be travelling to a polio endemic country prior to their preschool booster, in the form of Revaxis (dT-IPV). Otherwise, all children will be offered polio containing vaccine at the pre-school booster as per the national schedule.
- Hib <0.15 μg/ml
- MenC <8 SBA
- PCV ≥3 serotypes with <0.35 μg/ml

### 7 Intervention/Treatment of trial participants

#### 7.1 Study vaccines

Pregnant women will be randomised to receive REPEVAX or BOOSTRIX-IPV for protection against pertussis. In the UK, both vaccines are licensed for booster immunisation against diphtheria, tetanus, pertussis and poliomyelitis in persons from 3 years of age. REPEVAX has been used since October 2013 for maternal immunisation but will be replaced by BOOSTRIX-IPV from April 2014, as a result of the outcome of the national vaccine supply tender. In infants, all vaccines are licensed in the UK and routinely given as part of the recommended national immunisation schedule. The vaccines will therefore be used according to their SPCs or according to guidance by the Department of Health. The following vaccines will be used in this study:

**REPEVAX® ($DTa_5P$-IPV):** a combined vaccine that contains diphtheria toxoid (2 IU), tetanus toxoid (20 IU), pertussis (5 antigen component: 2.5μg PT, 5μg FHA, 3μg PRN, 5μg FIM 2 and 3) and polio (3 strains of inactivated virus – 40 D-antigen unit Types 1, 8 D-antigen unit Type 2, 32D-antigen unit Type 3). REPEVAX is currently recommended by the Department of Health for use in pregnancy

**BOOSTRIX-IPV® (dTap-IPV):** a combined vaccine that contains diphtheria toxoid (2 IU), tetanus toxoid (20 IU), pertussis (3 antigen component: 8μg PT, 8μg FHA and 2.5μg PRN) and polio (3 strains of inactivated virus – 40 D-antigen unit Types 1, 8 D-antigen unit Type 2, 32D-antigen unit Type 3).

BOOSTRIX-IPV is currently used in pregnancy in Flanders, Belgium and either REPEVAX® (known as Adacel) or BOOSTRIX may be administered in the United States. It is likely that BOOSTRIX-IPV will also be used in routine practice in the UK in the near future.

**Infanrix-IPV/Hib (DT $a_5$P-IPV-Hib):** a 5-in-1 combination vaccine that contains diphtheria, tetanus, pertussis (acellular, 3 antigen component), poliomyelitis (inactivated) and *Haemophilus influenzae* type b conjugate vaccine (adsorbed)

**NeisVac-C® (MCC-TT):** a meningococcal serogroup C conjugate vaccine with the meningococcal capsular polysaccharide covalently linked to tetanus toxoid

**Prevenar13® (PCV13; Pfizer Limited):** a 13-valent pneumococcal conjugate vaccine that protects against 13 different pneumococcal serotypes (1, 3, 4, 5, 6A, 6B, 7F, 9V, 14, 18C, 19A, 19F and 23F)

**Menitorix® (MCC-TT/Hib-TT; GlaxoSmithKline UK):** a combined *Haemophilus influenzae* type b (Hib) and meningococcal serogroup C conjugate vaccine where the capsular polysaccharides of both organisms are covalently linked to tetanus toxoid carriers

**Priorix® (MMR; GlaxoSmithKline UK):** a live attenuated vaccine that protects against measles, mumps and rubella.

**MMR®II (MMR; Sanofi Pasteur MSD)**: a live attenuated vaccine that protects against measles, mumps and rubella that may be used as an alternative to Priorix®, depending on which vaccine is received from the Department of Health by the GP surgery.

**Rotarix:** a live attenuated vaccine that is administered orally as a suspension in a pre-filled oral applicator. It protects against gastroenteritis caused by rotavirus.

Bexsero: a 4-component protein vaccine that protects against meningococcal serogroup B infections

#### 7.2 Vaccination procedures

At St. George's Hospital, the women will be vaccinated at the St. George's Vaccine Institute (SGVI) or St George's Clinical Research facility (CRF), the GP surgery, the family's home or the outpatients department, as preferred by the participants. In Hertfordshire and Gloucestershire, all vaccines will be administered by the Vaccine Research Nurses (VRNs) at the infants' GP surgery or at the participant's home if mutually agreeable to family and research nurse . Pregnant women will be vaccinated with either REPEVAX or BOOSTRIX-IPV according to their randomisation allocation and observed for 20 minutes. At visits 3, 4, 5, 7 and 8 infants will receive vaccines routinely given according to their Summary of Product Characteristics (SPCs) as part of the recommended national immunisation schedule.

The study team will ensure that the vaccines are maintained under appropriate storage conditions and record their batch numbers and expiry dates. Standard immunisation practices will be followed, according to the Green Book on Immunisation, 2013. GPs will be informed in writing of the vaccines administered to the women and infants (Appendix 0).

#### 7.3 Collection and posting of blood samples

For women, blood sampling will be via venepuncture; if blood collection has failed after two attempts at venepuncture, another operator may make one further attempt or they may be offered another appointment. Women will have the option of an anaesthetic cream or cryogenic spray to minimise discomfort. Up to 10ml of blood will be collected at each timepoint.

For infants under 3 months of age, oral sucrose solution (such as Sweet-Ease®) may be offered to minimise discomfort (subject to local practice/preference). If an infant is breast-fed, the mother will be asked if she wishes to breastfeed during the procedure to minimise discomfort. An anaesthetic cream (Denela, EMLA® or Ametop®) or cryogenic spray may be offered for infants one month of age or older. A minimum of 3 ml and a maximum of 5 ml of blood will be taken from each infant at each sample collection point. A smaller volume may be taken in infants weighing less than 5kg, see section 10.5. If blood collection has failed after two attempts at venepuncture, another operator may make one further attempt or they may be offered another appointment. Finger prick or heel prick may be attempted as a last resort if venepuncture proves difficult.

At St. George's Hospital visit 1 blood sample will be taken in the outpatients department, GP surgery, CRF or SGVI or at the woman's home prior to maternal vaccination. In Hertfordshire and Gloucestershire, visit 1 blood sample will be taken at the GP surgery or the woman's home prior to maternal vaccination. Visit 2 blood sample will be taken from the mother and the baby within 72 hours of birth. Participating women will be given a card to take to hospital when they are in labour with instructions to the hospital staff to collect and store a blood sample from the mother whilst in hospital and a cord blood sample from the baby at birth. Participating women will also be asked to contact the study team when they go to hospital. If a sample was not obtained from the woman or the baby, then Visit 2 will be arranged by the study team to obtain the blood sample(s), ideally within 72 hours of birth or as soon as possible after hospital discharge.

At St George's Hospital, subsequent visits will take place at SGVI or CRF, GP surgery, the outpatient department, or at the family's home. During subsequent visits in Hertfordshire and Gloucestershire, blood samples will be taken at the GP surgery or at the family's home, if more convenient for all concerned.

For all blood samples, the study team will stick an adhesive label bearing the participant's unique identification number and a bar code appropriate for the blood sample onto each sample tube. Samples will be appropriately packed and sent by Royal Mail special delivery (from Hertfordshire and St. George's Hospital) or Hayes Courier Service (from Gloucester) to the Immunoassay Laboratory of Porton Down, Public Health England (PHE Porton), in accordance with Royal Mail requirements, as follows:

**Royal Mail Special Delivery**

Samples collected are placed into a NOAX green topped tube containing an absorbent strip and then placed in an approved transport box (along with Sample Postage and Receipt Form CTD014). The box is then placed into a Royal Mail Special Delivery silver bag showing the "UN3373 biological substance, category B" identification diamond sticker. This is then posted via the local post office where a receipt will be issued to the sender. The samples are on a "before 1pm" next day delivery service and are trackable if a problem occurs.

**Hayes DX**

The samples are collected and placed into absorbent wraps within a large bio-bottle and then into bubble wrap to protect the samples during transit. This bio-bottle is then placed inside the Hayes Transport box (along with any associated paperwork) which is pre-printed with the "UN3373 biological substance, category B" identification diamond and has already had a bar-code attached by Hayes. A security seal is affixed to the outside of the box by the sender and they affix a sender (with emergency contact details) & addressee label. These are collected by Hayes DX for onward distribution by 1pm next day. This service is also trackable.

A sample postage form will be included with blood samples to document the movement of samples between sites and laboratories (Appendix). Logs of samples sent and received will be kept at PHE Colindale, to enable the identification of any lost or delayed samples and to provide a log of where samples are currently stored. Samples collected on Monday to Thursday will be posted daily;

samples collected on a Friday will be refrigerated over the weekend before being posted on Monday.

#### 7.4 Participant numbers and labelling of serum samples

Women and their infants will be assigned the same 4 digit number (1001 etc.). Numbers will be used sequentially in order of enrolment to the study within the blocks provided to each nurse or site. The first number will indicate the site of recruitment (Hertfordshire, Gloucestershire or St. George`s Hospital). This number, in addition to the protocol identification (e.g. iMAP2) and the blood sample letter (Blood MA, BA, etc.) will be recorded on all aliquots of serum as a barcode and text (e.g. iMAP2 1001 MA).

#### 7.5 Testing samples

Sera will be separated on arrival at the Porton laboratory, divided into aliquots and frozen at -80$^{o}$C or below. Frozen aliquots will be sent to: Virus Reference Department, PHE Colindale; Manchester PHE and the Institute of Child Health, London, at regular intervals for antibody measurement, following appropriate SOPs.

On completion of testing, samples will be archived at –80$^{o}$C or below at PHE Colindale if permission for this has been granted by the participants' parent/guardian; otherwise they will be destroyed. Samples will be tested for responses to the following, ***in order of priority*** as per the laboratory guidelines for this study:

- Pertussis (PT, PRN, FHA and then FIM 2 and 3) antibodies (PHE Porton)
- Anti-PRP (Hib antibody) IgG concentration (PHE Porton)
- Meningococcal serogroup C serum bactericidal antibody titres and serogroup C-specific IgG concentrations (PHE Manchester);
- Pneumococcal IgG concentration for the 13 serotypes contained in the vaccine and functional pneumococcal antibody studies (ICH, London);
- Tetanus antibodies (PHE Porton)
- Diphtheria antibodies (PHE Porton)
- Polio serotypes 1 2 and 3 (National Institute of Biological Standards and Control (NIBSC))
-

If sufficient serum is available functional pertussis immune studies and Men B responses will be performed (PHE Porton / St George's, University of London / PHE Manchester).

Any remaining sera will be stored with permission and may be used for other infection or vaccine related studies.

#### 7.6 Collection of safety data

Women who receive the pertussis vaccine in pregnancy will be observed for 20 minutes for any immediate reactions following vaccination. In infants, as all the vaccines administered are licensed, used according to their SPCs and offered to all infants as part of the national immunisation schedule, safety data using diaries will not be collected. At each visit, participating families will be asked about any adverse events that required a GP or hospital visit, and any such event will be recorded in the case report form (CRF).

##### 7.6.1 Definition of serious adverse events

Definitions of severe and serious adverse events are given in Appendix 0. Mothers will be asked to contact the study team as soon as possible if they or their baby have any severe/serious adverse event. A study clinician will judge the causal relationship between adverse event and vaccinations, after appropriate specialist consultation where necessary, as one of the following: probable, possible, or not related. Events will be graded by the study clinician as mild, moderate or severe.

##### 7.6.2 Reporting procedures for serious adverse events

A serious adverse event (SAE) occurring in any participant will be reported to the relevant REC and the Competent Authority (MHRA), where in the opinion of the Chief Investigator the event was 'related' (that is, it resulted from administration of any of the research procedures) and 'unexpected' (that is, the type of event is not listed in the summary of product characteristics as an expected occurrence).

Investigators, and/ or Research Nurses, will report any SAE immediately to the Sponsor. This will be coordinated by the Study Coordinator informing the PHE R&D Office (which acts of behalf of PHE as Sponsor). This is in conjunction with the UK Statutory instrument 2004 No 1031 part 5 and EC guidance document 2001/C 172/01 (CT-3) section 4.3 paragraph 29 which states that "Immediate reporting should allow the sponsor to take appropriate measures to address potential new risks in a clinical trial".

Reports of related and unexpected SAEs will be submitted within 15 days of the Chief Investigator becoming aware of the event (7 days in the event of a fatality), using the NRES report of serious adverse event form (see IRAS/NRES website). All SAEs will also be reported to the appropriate vaccine manufacturers where vaccine is obtained directly from them and not via the Department of Health routine stocks. All such events and any medication or other therapeutic measures used to treat the event will also be documented on the CRF, as well as in an update to the SAE form. Adequate documentation will be maintained to record that the appropriate notifications have been made. If required, a follow-up report including all new information obtained on the serious adverse event will be prepared and sent to the relevant authorities.

#### 7.7 Data Records

Study personnel will maintain their own records of all participants enrolled in the study under their care. CRFs will be provided by PHE Colindale. CRF packs will comprise a series of cards relating to each visit of the study. CRFs will be numbered and will indicate the participant's unique identification number. Also included will be a change of circumstances card for any participant whose name or address has changed during the course of their participation, and a completion card that will detail whether the protocol was completed or whether participants were withdrawn or lost to follow-up, with appropriate details given.

CRFs will be completed at each visit and sent to PHE Colindale. Information from CRFs will be entered at PHE Colindale into a study specific Access database. Data will be double entered by two independent administrators and verified electronically. As the data are being entered, the CRFs will be monitored for completion errors or omissions. When such a problem is identified the card will be photocopied and the field for correction marked. The photocopy will be sent to the responsible study nurse or doctor who will make the correction, crossing out any incorrect information with a single line, and will sign and date the change. On return of the photocopy to PHE Colindale, the database will be updated accordingly and the photocopy filed with the original CRF. Data from laboratories will be sent to the Clinical Trial Data Manager and imported into the database such that test results can be linked to clinical records.

#### 7.8 Withdrawal of Participants

Participants have the right to withdraw themselves and their infants from the study at any time and for any reason, without prejudice to his or her future medical care, and are not obliged to give their reasons for doing so. Whenever possible the reason for withdrawal will be recorded in the CRF. If the withdrawal is due to an adverse event, the study clinician will arrange for follow-up visits and/or telephone calls until the adverse event has resolved or stabilised. The study team may also withdraw a participant at any time in the interests of the mother's or the infant's health and well-being. In addition the participant may withdraw/be withdrawn for any of the following reasons:

- Administrative decision by the study team
- Ineligibility (either arising during the study or retrospective having been overlooked at screening)
- Significant protocol deviation
- Participant non-compliance with study requirements
- An adverse event that requires discontinuation of the study treatment or results in inability to continue to comply with study procedures

The results of all laboratory tests performed on blood samples of participants who withdraw or are withdrawn after providing written consent will be included in the data analyses unless specifically requested by the family not to do so.

#### 7.9 Duration of the study

It is hoped that recruitment to the study will be completed in a 12 month timeframe. However, as with all vaccine studies, the recruitment rate depends on parental attitudes and is largely unpredictable. The schedule of vaccination and sample collection may span from 28 weeks gestation to the infant reaching 13 months of age, equating to around 18 months for each mother/child pair. The study involves eight visits for women who receive one of the pertussis containing vaccines in pregnancy (women who elect not to receive a pertussis vaccine in pregnancy will only require study visits 3 - 8 and the total duration will be up to 13 months).

#### 7.10 Definition of the end of the trial

The end of the trial is the date of reporting of the final blood test result.

#### 7.11 Expenses and Payments

Because study personnel will meet the family at their home or their local surgery in most instances, it is not expected that the participants will incur any costs.

### 8 Statistical Methods

Full details will be given in the statistical analysis plan produced by the Immunisation Department Statistician.

#### 8.1 PRIMARY END-POINT

- **Infants at 5 months of age whose mothers received either REPEVAX or BOOSTRIX-IPV**: fold-difference in anti-PT IgG GMC

#### 8.2 SECONDARY END-POINTS

- **Infants at birth whose mothers received either REPEVAX or BOOSTRIX-IPV**: Concentration (GMC) of IgG to pertussis antigens (PT, PRN, FHA and FIM 2 and 3), tetanus toxoid and diphtheria toxoid
- **Infants at 2, 5 and 13 months whose mothers received either REPEVAX, BOOSTRIX-IPV or no pertussis-containing vaccine**: Concentration (GMC) of IgG to pertussis antigens (PT, PRN, FHA and FIM 2 and 3); anti-PRP IgG [Hib antigen] GMC and proportion of infants with concentrations ≥ 0.15 and ≥ 1.0; anti-tetanus toxoid IgG GMC; anti-diphtheria toxoid IgG GMC; meningococcal serogroup C serum bactericidal antibody titres (GMT) and proportion of infants with titres ≥ 8 and ≥128; meningococcal serogroup C-specific IgG GMC; ; 13 serotype-specific pneumococcal IgG GMC and functional pneumococcal antibody levels.
- **Mothers pre-vaccination and at delivery who received REPEVAX or BOOSTRIX-IPV**: Concentration (GMC) of IgG to pertussis antigens (PT, PRN, FHA and FIM 2 and 3), tetanus toxoid and diphtheria toxoid;.
- **Mothers who received REPEVAX or BOOSTRIX-IPV**: Placental transfer ratio of IgG to pertussis antigens (PT, PRN, FHA and FIM 2 and 3), tetanus toxoid ,diphtheria toxoid;

- **Mothers who received either REPEVAX, BOOSTRIX-IPV or no pertussis-containing vaccine**: Persistence of pertussis IgG (PT, PRN, FHA and FIM 2 and 3) in mothers at 13 months after delivery measured as the geometric mean ratio of concentrations at 13months versus concentrations at delivery.

##### 8.3. EXPLORATORY ENDPOINTS

Comparisons in serotype-specific polio antibody levels (GMTs and percentages with titres ≥1 in 8 by microneutralization) will be made between unvaccinated mothers and their infants and vaccinated mothers and their infants with data for the latter combined across the two TdaP/IPV vaccines used for maternal immunisation

##### 8.4 The number of participants

Based on sample size calculations below, it is proposed that a convenience sample of 65 (+/-10) mother-infant pairs will be recruited into each of the two groups that received a pertussis vaccine in pregnancy, with a control group of 50 (+/-10) women and their infants. Up to 200 mother-infant pairs will be recruited to take account of mother-infant pairs lost to follow-up.

The sample size calculation is based on data from the recent P13UK study performed by PHE. Based on these results, the standard deviation of the post-primary vaccination GMCs (log-10 scale) was:

PT = 0.28

Hib =0.76

MenC SBA = 0.50

Tet =0.37

Based on these results the detectable fold differences (5% significance, 80% power) between the two vaccinated study arms are in table 1. A range of sample sizes from 50 to 125 are given in the table to show how precision changes with sample size.

| Number of participants | **PT** | **Hib** | **SBA** | **Tet** |
|---|---|---|---|---|
| 50 | 1.45 | 2.67 | 1.92 | 1.61 |
| 65 | 1.38 | 2.37 | 1.77 | 1.52 |
| 80 | 1.33 | 2.18 | 1.67 | 1.46 |
| 100 | 1.30 | 2.00 | 1.58 | 1.39 |
| 125 | 1.26 | 1.86 | 1.51 | 1.35 |

Table1. Summary of detectable fold differences (5% significance, 80% power) for different numbers of participants

*For example, With a sample size of N=65 in each group of vaccinated women, we can detect a difference between a geometric mean PT level of 20 in one group and 28 in the other group.*

In the P13UK study, Hib GMC and MenC GMT were 3-4-fold lower for the CRM/TT group compared with the second lowest group (CRM/CRM for Hib and TT/TT for MenC). Table 1 summarises the 95% CIs around various observed percentages in each group (these could be either percentages with reactions (0% to 20%) or responding (70% to 100%)). Table 2 shows the percentages that are detectable as different in one group for various percentages in the other group (80% power, 5% significance).

| | **SAMPLE SIZE** | | | |
|---|---|---|---|---|
| **Percentage** | **50** | **65** | **80** | **100** |
| 0 | 0 to 7.1 | 0 to 5.5 | 0 to 4.5 | 0 to 3.6 |
| 10 | 3.3 to 21.8 | 3.9 to 20.0 | 4.4 to 18.8 | 4.9 to 17.6 |
| 20 | 10.0 to 33.7 | 11.1 to 31.8 | 11.9 to 30.4 | 12.7 to 29.2 |
| 70 | 55.4 to 82.1 | 57.4 to 80.7 | 58.7 to 79.7 | 60.0 to 78.8 |
| 85 | 72.1 to 93.5 | 74.0 to 92.6 | 75.3 to 92.0 | 76.5 to 91.4 |
| 90 | 78.2 to 96.7 | 80.0 to 96.1 | 81.2 to 95.6 | 82.4 to 95.5 |
| 95 | 84.8 to 99.2 | 86.6 to 98.9 | 87.7 to 98.6 | 88.7 to 98.4 |
| 100 | 92.9 to 100 | 94.5 to 100 | 95.5 to 100 | 96.4 to 100 |

Table 1. Summary of 95% CIs around various observed percentages in each group

*For example, with a sample size of 65, 95% CI around an observed percentage of 85% would be 74.0 to 92.6.*

| | Sample Size per group | | | |
|---|---|---|---|---|
| **Percentage** | 50 | 65 | 80 | 100 |
| 0 | -, 17.7 | -,13.8 | -,11.4 | -,9.3 |
| 10 | -, 35.1 | -,31.2 | -,28.6 | 0.3,26.1 |
| 20 | 1.1, 48.2 | 3.0,44.3 | 4.4,41.5 | 5.8,38.9 |
| 70 | 40.5, 93.4 | 44.4,90.8 | 47.1,88.9 | 49.7,87.1 |
| 85 | 58.1, - | 62.0,99.5 | 64.7,98.4 | 67.2,97.2 |
| 90 | 64.9, - | 68.8,- | 71.4,- | 73.9,99.7 |
| 95 | 72.7, - | 76.5,- | 80.0,- | 81.3,- |
| 100 | 82.3, - | 86.1,- | 88.6,- | 90.7,- |

Table 2. Summary of percentages that are detectable as different in one group for various percentages in the other group (80% power, 5% significance).

*For example, with a sample size of 65 then if one group has an expected 95% with protective titres then there is 80% power to detect this as different from 76.5% in the other group. It is not possible to detect 95% as different from 100% which is why a – is given.*

#### 8.5 Data analysis and presentation

Data analysis will be undertaken by the PHE Colindale Statistician and members of the study team where appropriate. The results of the study will be submitted to the Joint Committee for Vaccination and Immunisation (JCVI), the Department of Health and published in a peer-reviewed journal. For the analysis logged titres will compared between study arms at each time point using normal errors regression and t-tests or a non-parametric test (Kruskal Wallis) if appropriate. Proportions above titre thresholds will be compared using Fisher's exact test. Geometric mean titres, concentrations and fold differences, as well as proportions above thresholds will also be calculated with 95% confidence intervals. For persistence geometric mean fold changes will be calculated with 95% CI. For the analysis logged titres will compared between study arms at each time point using normal errors regression and t-tests or a non-parametric test (Kruskal Wallis) if appropriate. Proportions above titre thresholds will be compared using Fisher's exact test. Geometric mean titres, concentrations and fold differences, as well as proportions above thresholds will also be calculated with 95% confidence intervals. For persistence geometric mean fold changes will be calculated with 95% CI. Further details will be in the study specific statistical analysis plan.

### 9 QUALITY CONTROL & QUALITY ASSURANCE PROCEDURES

Data will be evaluated for compliance with the protocol and accuracy in relation to source documents. Regular monitoring will be performed according to ICH GCP to verify that the clinical trial is conducted and data are generated, documented and reported in compliance with the protocol, GCP and the applicable regulatory requirements. All sites will provide direct access to all trial related source data/documents and reports for the purpose of monitoring and auditing by the Sponsor and inspection by local and regulatory authorities.

### 10 ETHICS AND REGULATORY AUTHORITIES

#### 10.1 Declarations

##### 10.1.1 Clinical trials regulations

This protocol will be conducted in accordance with the Medicines for Human Use (Clinical Trials) Regulations 2004 - as detailed in statutory instrument SI 2004 No1031, and also SI 2006 No. 1928.

##### 10.1.2 Declaration of Helsinki

The Investigators will ensure that this study is conducted in full conformity with the current revision of the Declaration of Helsinki (last amended October 2008).

##### 10.1.3 ICH Guidelines of Good Clinical Practice

The Investigator will ensure that this study is conducted in full conformity with relevant regulations and with the ICH Guidelines for Good Clinical Practice (CPMP/ICH/135/95) July 1996.

##### 10.1.4 Laboratories

Laboratorieswill follow “MHRA Good Clinical Practice guidance on the maintenance of regulatory compliance in laboratories that perform analysis or evaluation of clinical trial samples”, Issue 1, July 2009 or will have CPA accreditation for the tests being performed where the test result comprises a secondary objective.

#### 10.2 Registrations

PHE has obtained a Eudract registration number for the trial, and the study will also be registered in an appropriate publically accessible clinical trials registry.

#### 10.3 Approvals

##### 10.3.1 Ethics Approvals

All relevant documents, including the protocol, informed consent forms, and parent information sheets will be submitted to a Research Ethics Committee (REC).

Where amendments are needed to the originally approved documents, the Investigator will submit and, where necessary, obtain approval from the REC for amendments in accordance with applicable regulations and requirements.

##### 10.3.2 NHS Approvals

The appropriate NHS R&D approvals will be sought for the inclusion of patients locally.

The study team has established links with the NIHR Research Networks in all sites. This study will be put forward for appropriate Research Network adoption and applicable resources.

##### 10.3.3 MHRA

A Clinical Trial Authorisation (CTA) will be applied for and obtained from the MHRA prior to the start of the study.

#### 10.4 Participant Confidentiality

Personal data collected for the purposes of this study may include name, date of birth, address as well as the blood test results and any relevant medical information required to analyse the vaccine immune responses. The only people with access to this information will be PHE and SGVI staff, Department of Health or regulatory authorities who may wish to check the study is being carried out according to appropriate guidelines. Every effort will be made to protect the participants' identity. Blood samples will only be identified only by a bar-coded sticker with the participant's study number. Data will only be used for the purposes of this study, stored in secure PHE and SGUL facilities and only accessed by trial staff and authorised personnel and destroyed after an appropriate time period. The study will comply with the Data Protection Act, which requires data to be anonymised as soon as it is practical to do so.

#### 10.5 Other Ethical Considerations

Venepuncture may be uncomfortable, but every effort will be made to reduce any discomfort associated with blood sampling. All procedures will be performed by fully trained nurses or doctors who are experienced in vaccinating and obtaining blood samples from adults and young infants.

The volume of blood collected from infants will be within the guidance given by the European Medicines Agency (http://ec.europa.eu/health/files/eudralex/vol-10/ethical_considerations_en.pdf): The study-related blood volume collected will not exceed 3% of the total blood volume in any 4 week period or 1% at any time. In practice this means that any peripheral blood sampling of an average newborn infant will not exceed 3mls (based on weight of 3.5kg and blood volume of 90mls/kg). For smaller infants, the volume of blood collected at birth will be adjusted according to weight, such that a maximum of 2mls will be collected from a 2.5 kg baby and 1ml from a 1.5kg baby. It is anticipated that cord blood will be collected in most infants therefore a peripheral blood sample at birth would not be required.

At visit 2, the average infant will weigh around 5kg and therefore the maximum blood volume collected will be 4.5mls. This will be adjusted accordingly for smaller infants.

At subsequent visits, all infants would be expected to be of sufficient size for a maximum of 5mls to be safely collected. However in smaller infants this will be adjusted accordingly.

Any participant found to have a low Hib, pneumococcal or MenC in the sample taken at about 13 months of age (following the boosters given at about 12 months of age) will be contacted to discuss the results and to recommend further dose(s) of vaccine, which will be administered by the study team. Any participant found to have a low polio response AND who will be visiting a polio endemic country prior to their pre-school booster, will be offered an extra dose in the form of dT-IPV, currently available as Revaxis.

### 11 SPONSORSHIP AND INDEMNITY

The study is sponsored by Public Health England (PHE). In accordance with the 2004 EU Clinical Trial Directive, indemnity arrangements will be made by the Sponsor. As the Sponsor for this study will be PHE, indemnity for study staff conduct will be provided under the Crown Indemnity Scheme, with provision for cover for any negligent harm caused by staff as a result of study participation and where justified, an ex gratia payment for any non-negligent harm.

### 12 PUBLICATION POLICY

The Investigators will be involved in reviewing drafts of manuscripts, abstracts and any other publications arising from the study. Authorship will be determined in accordance with the ICMJE guidelines and other contributors will be acknowledged.

### 13 REFERENCES

ACIP. 2011. Updated Recommendations for Use of Tetanus Toxoid, Reduced Diphtheria Toxoid and Acellular Pertussis Vaccine (Tdap) in Pregnant Women and Persons Who Have or Anticipate Having

Close Contact with an infant Aged <12 months – Advisory Committee on Immunization Practices (ACIP), 2011. MMWR October 2011; 60(41): 1424-1426.

Centers for Disease Control and Prevention (CDC). Updated recommendations for use of tetanus toxoid, reduced diphtheria toxoid and acellular pertussis vaccine (Tdap) in pregnant women and persons who have or anticipate having close contact with an infant aged <12 months --- Advisory Committee on Immunization Practices (ACIP), 2011.

Department of Health. Information about whooping cough vaccination programme for pregnant women. 28 September 2010. Available at: http://www.dh.gov.uk/health/2012/09/whooping-cough-information/

Department of Health. Whooping cough vaccination programme for Pregnancy Women. 2012 : Available at: https://www.gov.uk/government/publications/whooping-cough-vaccination-programme-for-pregnant-women

Department of Health, 2013. Continuation of temporary programme of pertussis (whooping cough) vaccination of pregnant women. Available at: htttps://www.gov.uk/government/uploads/system/uploads/attachment_data/file/197839/130510_Pertussis_continuation_letter_FINAL.pdf

Englund JA, Anderson EL, Reed GF, Decker MD, Edwards KM, Pichichero ME, Steinhoff MC, Rennels MB, Deforest A, Meade BD. The effect of maternal antibody on the serologic response and the incidence of adverse reactions after primary immunization with acellular and whole-cell pertussis vaccines combined with diphtheria and tetanus toxoids. Pediatrics. 1995 Sep;96(3 Pt 2):580-4.

de Greeff SC, Mooi FR, Westerhof A, Verbakel JM, Peeters MF, Heuvelman CJ, Notermans DW, Elvers LH, Schellekens JF, de Melker HE. Pertussis disease burden in the household: how to protect young infants. Clin Infect Dis. 2010 May 15;50(10):1339-45.

Halasa, N. B., O'Shea, A., Shi, J. R., LaFleur, B. J., & Edwards, K. M. (2008). Poor immune responses to a birth dose of diphtheria, tetanus, and acellular pertussis vaccine. The Journal of pediatrics, 153(3), 327–332.

Halperin BA, Morris A, MacKinnon-Cameron D et al. Kineticsa of the antibody response to tetanus-diphtherias-acellular pertussis vaccine (Tdap) in women of child bearing age and post partum women. Clin Infect Dis 2011; 53:885-892.

Health Protection Agency, 2012. Pertussis notifications and deaths. PHE website. Available at. http://www.PHE.org.uk/webc/PHEwebFile/PHEweb_C/1317133571994

Health Protection News, 2012. Confirmed pertussis in England and Wales continues to increase. Available at: http://www.hpa.org.uk/hpr/archives/2012/news1512.htm

Health Protection Reports, 2012. Laboratory confirmed cases reported to the enhanced pertussis surveillance programme in 2011. Health Protection Report vol 6(8); Feb 2012. Available at http://www.PHE.org.uk/hpr/archives/2012/hpr0812.pdf

Gall SA, Myers J, Pichichero M. Maternal immunization with tetanus-diphtheria-pertussis vaccine: effect on maternal and neonatal serum antibody levels. Am J Obstet Gynecol. 2011 Apr;204(4):334.e1-5. Epub 2011 Jan 26.

Knuf, M., Schmitt, H.-J., Wolter, J., Schuerman, L., Jacquet, J.-M., Kieninger, D., et al. (2008). Neonatal vaccination with an acellular pertussis vaccine accelerates the acquisition of pertussis antibodies in infants. The Journal of pediatrics, 152(5), 655–60– 660.e1.

Leuridan E, Hens N, Peeters N, de Witte L, Van der Meeren O, Van Damme P. Effect of a prepregnancy pertussis booster dose on maternal antibody titers in young infants. Pediatr Infect Dis J. 2011 Jul;30(7):608-10.

Mulholland, K., Suara, R. O., Siber, G., Roberton, D., Jaffar, S., N'Jie, J., et al. (1996). Maternal Immunization With Haemophilus influenzae Type b Polysaccharide—Tetanus Protein Conjugate Vaccine in The Gambia. The Journal of the American Medical Association, 275(15), 1182–1188.

Van Savage J, Decker MD, Edwards KM, Sell SH, Karzon DT. Natural history of pertussis antibody in the infant and effect on vaccine response. J Infect Dis. 1990 Mar;161(3):487-92

Safety and immunogenicity of Tdap vaccine in healthy pregnant women, safety in their neonates, and effect of maternal immunization on infant immune responses to dTap Vaccine. Available at: www.clinicaltrials.gov. Sponsor: National Institute of Allergy and Infectious Diseases (NIAID). ClinicalTrials.gov Identifier: NCT00707148. Accessed 29 December 2010.

Immunization of women with diphtheria and tetanus toxoids combined with acellular pertussis (Tdap) during the mid third trimester of pregnancy: An evaluation of the potential for immunological protection for the neonate. Available at: www.clinicaltrials.gov. (Sponsor: Dalhousie University; collaborators: IWK Health Centre, Sanofi Pasteur.). ClinicalTrials.gov Identifier: NCT00553228. Accessed 29 December 2010.

### Appendix: associated documents

1 INFORMATION FOR PROSPECTIVE PARTICIPANTS: COVERING LETTERS, PRE-INFORMATION LEAFLETS AND PARTICIPANT INFORMATION LEAFLETS

1.1 Covering letter 1 for use at St George's Hospital in the antenatal clinics (CovLetSGH1) 44

**3 CORRESPONDENCE WITH GP**

**3.1 Letter to GP notifying of participation in study: randomised women ... 71**

**3.2 Letter to GP notifying of participation in study: unvaccinated women ... 72**

**3.3 Letter to GP notifying of vaccination of women ... 73**

**4 ADVERSE EVENTS ... 74**

**5 PARTICIPATING ORGANISATIONS AND STUDY PERSONNEL ... 76**

### 1 Information for prospective participants: Covering letters, pre-information leaflets and participant information leaflets

#### 1.1 Covering letter 1 for use at St George's Hospital in the antenatal clinics (CovLetSGH1)

Public Health England

St George's Healthcare NHS Trust

St George's University of London

Dear Parent,

**Re: Clinical study being carried out at St George's Hospital - immunising Mums Against Pertussis (iMAP2)**

St George's Hospital and St George's University are working with Public Health England (PHE) on a study to assess two whooping cough (pertussis) vaccines in pregnancy. PHE is part of the Department of Health and has the remit of protecting the health and well-being of the population.

We are giving you this letter as you are currently pregnant and will be offered the whooping cough vaccine from 28 weeks of pregnancy under the routine national vaccination schedule. The enclosed leaflet gives a brief summary about the study and tells you how you can receive more information.

Please note that neither St George's University nor PHE has your name or address details and will only contact you if you request further information.

If you do not want a member of the study team to contact you, you need do nothing further and please ignore any reminder card that we may send you. This will not affect your or your baby's routine medical care. However, we would appreciate knowing the reason why you do not wish to partake in this study if at all possible.

Yours sincerely

Representative of Maternity Services, St George's Hospital: name and signature

#### 1.2 Covering letter 1 for use by GPs in South London for women in the antenatal period (CovLetSGH-GP1)

Public Health England St George's Healthcare NHS Trust St George's University of London

Dear Parent,

**Re: Clinical study being carried out at St George's Hospital - immunising Mums Against Pertussis (iMAP2)**

We are working with St George's University and Public Health England (PHE) on a study to assess two whooping cough (pertussis) vaccines that are currently used in pregnancy. PHE is part of the Department of Health and has the remit of protecting the health and well-being of the population.

We are sending you this letter as you are currently pregnant and will be offered the whooping cough vaccine from 28 weeks of pregnancy under the routine national immunisation schedule. The enclosed leaflet gives a brief summary about the study and tells you how you can receive more information.

Please note that neither St George's University nor PHE has your name or address details and will only contact you if you request further information.

If you do not want a member of the study team to contact you, you need do nothing further and please ignore any reminder card that we may send you. This will not affect your or your baby's routine medical care. However, we would appreciate knowing the reason why you do not wish to partake in this study if at all possible.

Yours sincerely

GP/representative signature & name

1.3 Covering letter 1 for use at PHE sites in the antenatal period (CovLetPHE1)

Public Health England

St George's University of London

Dear Parent,

**Re: Clinical study - immunising Mums Against Pertussis (iMAP2)**

We are working with Public Health England (PHE) and St George's University on a study to assess two whooping cough (pertussis) vaccines in pregnancy. PHE is part of the Department of Health and has the remit of protecting the health and well-being of the population.

We are sending you this letter as you are currently pregnant and will be offered the whooping cough vaccine from 28 weeks of pregnancy under the routine national immunisation schedule. The enclosed leaflet gives a brief summary about the study and tells you how you can receive more information.

Please note that the PHE does not have your name or address details and will only contact you if you request further information.

If you do not want a Vaccine Research Nurse based at our practice to contact you, you need do nothing further and please ignore any reminder card that we may send you. This will not affect your or your baby's routine medical care. However, we would appreciate knowing the reason why you do not wish to partake in this study if at all possible.

Yours sincerely

GP/representative signature & name

##### 1.4 Pre-information leaflet 1 for use at all sites in the antenatal period (PreInfo1)

**Immunising Mums Against Pertussis (iMAP2)**

- Public Health England (PHE) and St George's University are working with GP surgeries to compare two whooping cough (pertussis) vaccines given in pregnancy, on behalf of the Department of Health
- There is a national outbreak of whooping cough and young babies are at highest risk of getting whooping cough and having more severe disease. Vaccinating pregnant women helps protect them against infection because antibodies cross the placenta from mother to baby.
- We would give you a whooping cough vaccine and your baby all their routine vaccines. By taking small blood samples we will see how you and your baby respond to the vaccines.
- If your baby has low antibody levels against certain infections, we will arrange for them to have a booster dose of vaccine to make sure they are well protected.

**If you would like further information please contact the study team by either:**

(i) Completing the reply slip at the bottom of this leaflet and returning it in the envelope provided.
(ii) Texting iMAP2 plus your name and GP surgery to the mobile number XXX

Thank You

✂-----------------------------------------------------------------------------------------------------------------------------

I would like to receive further information about this study and I am happy for the study team to contact me.

Name: ______________________________________________ Due Date:__________

Address:______________________________________________________________

______________________________ Postcode:____________________________

Tel No:_________________________ Mobile no.______________________

Best time to contact me: ____________ GP Surgery I attend: ____________________

For office reference: iMAP2 (Randomised)

1.5 Covering letter for participant information leaflet in antenatal period 1: all GP sites (CovLetPILSGH)

Public Health England

St George's Healthcare NHS Trust

St George's University of London

St George's Vaccine Institute

www.vaccine.ac.uk

0208 725 3887

mobile number: XXXXXX

Dear Parent

**immunising Mums Against Pertussis (iMAP2)**

Thank you for requesting information about the study of whooping cough (pertussis) vaccines in pregnancy. Please find enclosed a leaflet that gives full details of the study and what would be involved should you wish to enrol yourself and your baby.

I will try to contact you by telephone shortly to see if you would like to discuss any aspect of the study or have any queries. Taking part in the study is entirely voluntary. If you decide not to participate, it will in no way affect your or your baby's routine care and their immunisations will be offered by your surgery in the usual way.

If you would like to take part in the study you can also contact me on the contact details above (if you prefer to decline you need take no action). I look forward to hearing from you and thank you for your time and consideration.

Yours sincerely

iMAP2 study team

1.6 Covering letter for participant information leaflet in antenatal period 1: PHE sites (CovLetPILPHE)

Public Health England

St George's University of London

Dear Parent

**Immunising Mums Against Pertussis (iMAP2)**

I am the Vaccine Research Nurse at your doctor's surgery. Thank you for requesting information about the study of whooping cough (pertussis) vaccines taking place at the surgery. Please find enclosed a leaflet that gives full details of the study and what would be involved should you wish to enrol yourself and your baby.

I will try to contact you by telephone shortly to see if you would like to discuss any aspect of the study or have any queries. Taking part in the study is entirely voluntary. If you decide not to participate, it will in no way affect your or your baby's routine care and their immunisations will be offered by your surgery in the usual way.

If you would like to take part in the study you can also call me on the telephone numbers listed at the bottom of this page. (If you prefer to decline you need take no action). I look forward to hearing from you and thank you for your time and consideration.

Yours sincerely

Vaccine Research Nurse

[VRN CONTACT DETAILS]

#### 1.7 Participant information leaflet 1 for use at all sites in the antenatal period (PIL1)

Public Health England

St George's Healthcare NHS Trust

St George's University of London

**immunising Mums Against Pertussis (iMAP2)**

**Full title: A head-to-head randomised controlled trial comparing two pertussis-containing vaccines in pregnancy and vaccine responses in UK mothers and their infants**

Public Health England and St George's University are conducting a study on behalf of the Department of Health assessing two whooping cough (pertussis) vaccines used in pregnancy. You have been contacted because you are pregnant and will be offered the whooping cough vaccine from 28 weeks of pregnancy, in line with national guidelines.

Vaccines activate the immune system to make antibodies, which are the components in the blood that protect against infections. We will measure the levels of antibody to the whooping cough vaccine given in pregnancy in mothers and babies and also look at the babies' response to the routine infant vaccines given at 2, 3 and 4 months of age. We would like to invite your family to take part in this study.

The study will involve nearly 130 women and their babies, with Mums receiving a whooping cough vaccine in pregnancy. We will also enrol 50 women who decided not to receive a whooping cough vaccine in pregnancy to take part, these women and their infants will be invited to take part after the baby has been born.

Before you decide if you would like you and your baby to join this study, you need to understand the reasons why this study is being done and what it would involve. Please take the time to read the following information carefully and talk to others about the study if you wish.

**BOX FOR ST GEORGE'S**
**In summary:**

- The study involves taking three blood samples from mums and three (or four if no cord blood is collected) from babies to check the amount of antibody made in response to vaccinations given under the routine UK schedule.
- If the blood test shows low antibody levels against Meningitis C, pneumococcal or Hib vaccines, the study team will contact you to recommend an extra dose of vaccine for your baby. We will also offer an extra dose of polio vaccine if your baby has a low response and if you are planning to visit certain parts of the world where the risk of infection is highest, your nurse will discuss this with you.

**BOX FOR ST PHE sites (Herts and Glouc)**
**In summary:**

- The study involves taking three blood samples from mums and four from babies to check the amount of antibody made in response to vaccinations given under the routine UK schedule.
- If the blood test shows low antibody levels against Meningitis C, pneumococcal or Hib vaccines, the study team will contact you to recommend an extra dose of vaccine for your baby. We will also offer an extra dose of polio vaccine if your baby has a low response and if you are planning to visit certain parts of the world where the risk of infection is highest, your nurse will discuss this with you.

**What is the purpose of the study?**
In September 2012, the UK Department of Health introduced an immunisation programme for pregnant women with whooping cough vaccination at 28-38 weeks of pregnancy. This recommendation was made following a marked rise in whooping cough cases across all age groups, but particularly in young infants who are at increased risk of severe disease, which may result in hospitalisation and death.

Vaccinating women during pregnancy allows their baby to be protected in the first weeks of their life, when they are at greatest risk, as the antibodies from the mum cross the placenta to the baby. Both of the whooping cough vaccines also give protection against tetanus, diphtheria and polio across the placenta. Babies are vaccinated themselves against these infections at 2, 3 and 4 months of age. This study will assess the antibodies you and your baby have to the vaccinations and will be important both for you to be reassured they have protection as well as helping the Department of Health continue to ensure the best protection is offered to our population.

**Why am I and my baby being invited to take part?**
We are inviting all pregnant women being looked after at St George's Hospital or who are registered with participating GP surgeries in Hertfordshire, Gloucestershire and South London. We will enrol up to 200 pregnant women and then their babies over the next few months. We will include women who chose to receive a whooping cough vaccine in pregnancy as well as those who decided not to have whooping cough vaccine in pregnancy.

**Do we have to take part?**
No, it is up to you to decide whether you would like to take part. If you agree you and your baby will take part, you will be asked to sign a consent form. Participation is entirely voluntary and you can withdraw from the study at any time without giving a reason and this would not affect either your or your baby's routine care. If you do not wish to participate or if you withdraw during the study, you will both be offered your and your baby's routine immunisations through the GP surgery. If a blood sample has already been taken, you will need to decide if you want the sample to be destroyed, in which case you will need to inform us in writing. If the sample has already been tested, we will send you and your GP the results and discuss any further vaccinations advised with you.

**What will happen to me and my baby if we take part?**
This study involves three blood samples from you: before your whooping cough vaccine, around the time you give birth and at one year after your baby is born. These will allow us to measure your

antibody levels before the vaccine, allow us to estimate the amount of antibody transferred to your baby and to see how long the antibodies last after the vaccine.

PARAGRAPH FOR ST GEORGE'S
Most babies in the study will have three blood samples taken: one before the first of the routine infant vaccines (these are the vaccines given at 2, 3 and 4 months of age), one after the last routine vaccination and then one after the booster vaccines (these are given around their 1st birthday). We will also collect a sample of blood from the umbilical cord after your baby is born. This is done after the cord has been clamped and cut and taken from the part of the umbilical cord that is no longer attached to your baby. Where it has not been possible to collect this cord blood, we will ask you for a blood sample from your baby. For these few babies, it would mean four blood tests in total over a year.

PARAGRAPH FOR HERTS/ GLOUC
Most babies in the study will have four blood samples taken: one in the first week after they are born, one before the first of the routine infant vaccines (these are the vaccines given at 2, 3 and 4 months of age), one after the last routine vaccination and then one after the booster vaccines (these are given around their 1st birthday).

The blood samples from your baby will allow us to estimate the level of protective antibody your baby gets from you through the placenta and how well they produce their own antibodies after their vaccines. We will offer ways to reduce any discomfort during the blood samples for you and your baby, such as a cream to numb the skin.

The timing of the vaccinations and blood tests is shown in the table below. The first appointment should last around an hour and all following appointments about 45 minutes.

If you agree for you and your baby to take part, a member of the study team will arrange an appointment to meet you at your convenience to answer any further questions that you may have, check your health and take a small sample (about 2 teaspoons) of blood. In order to reduce any discomfort, the nurse will offer you some local anaesthetic cream or cold spray to numb the skin before taking the blood sample. We will give you one of the whooping cough vaccines.

PARAGRAPH FOR ST GEORGE'S ONLY
When you go into labour, please take the pack we give you to the hospital so that the cord blood can be collected after your baby has been born, or give to the midwife if you opted for a home birth.

Please let us know when you are in labour or as soon as possible after your baby has been born so we can arrange your baby's first vaccination appointment with you.

The study team will give your baby all of the vaccinations according to the routine national schedule, i.e. what your baby would otherwise be given at your GP surgery, and will take the blood samples. The amount of blood collected from your baby will be less than one teaspoon full.
In South London all the study visits will take place at your home or at St George's Hospital. In Hertfordshire and Gloucester the Vaccine Research Nurse based will meet you at your GP surgery or at home as appropriate.

If the study team is unable to obtain a blood sample from your baby, you will be given the option to re-schedule the appointment or miss that sample.

We will let your GP know that you and your baby are taking part in the study. We will write to you and your GP with the results of the blood tests. If your baby does not develop enough antibodies against the Meningitis C, pneumococcal or Hib vaccines, the study team will contact you to offer your baby an extra dose of vaccine. We will also offer an extra dose of polio vaccine if your baby has a low response and if you are planning to visit certain parts of the world where the risk of infection is highest, your nurse will discuss this with you.

The diagram below summarises what will happen in the study – visits are shown according to the baby's age

| Visit | Pregnancy | Birth | 2 months | 3 months | 4 months | 5 months | 12 months | 13 months |
|---|---|---|---|---|---|---|---|---|
| **Mother** | REPEVAX<br>or<br>BOOSTRIX-IPV<br><br>Blood test | Blood test | | | | | | Blood test |
| **Baby** | | Blood test or cord blood sample | Routine infant vaccinations<br><br>Blood test | Routine infant vaccinations | Routine infant vaccinations | Blood test | Routine booster vaccinations | Routine booster vaccinations<br><br>Blood test |

Routine vaccinations are set out in the "Red Book" which you will have been given by your midwife. The schedule is available at: http://www.nhs.uk/Tools/Pages/NHSvaccinationplanner.aspx#close

**What are the alternatives?**

If you do not wish to take part in this study, then you do not need to do anything. Your GP surgery will contact you for the routine vaccinations at the appropriate times.

**What are the possible disadvantages and risks as well as benefits of taking part?**

This study will involve blood tests that are not part of routine care. However, your baby will have the advantage of being offered an extra dose of vaccine if antibodies against polio, Hib, pneumococcal or Meningitis C are found to be low. We will also provide the routine vaccines at a time and a place that are convenient for you. There is also a benefit to the community, as our results will be used to inform future vaccination policy by the Department of Health.

Both vaccines have been used, or are in current use, for pregnant women in the UK.. Although the vaccines are not licensed for pregnant women this is because pregnant women are routinely excluded from clinical trials, and not because of any specific safety concerns or evidence of harm in pregnancy. The JCVI which is the body that advises the Department of Health on vaccine matters has stated that "There is no evidence of risk to pregnancy or the infant with inactivated viral or bacterial vaccines or toxoids such as those against diphtheria, tetanus, polio and pertussis (whooping cough)".

**Why are you comparing two different whooping cough vaccines?**

Both of the vaccines protect against the same diseases: whooping cough as well as diphtheria, tetanus and polio but we want to ensure that both of these vaccines protect infants equally well.

**What are the side-effects when taking part?**

The blood test may be uncomfortable but will be performed by experienced nurses or doctors. As with all vaccines, there may be some discomfort, redness and mild swelling where the injection is given. We do not anticipate any other side-effects.

**What happens when the study stops?**
We will write to you with the results of your baby's blood tests. If your baby does not have enough antibodies against the polio, Meningitis C, pneumococcal or Hib vaccines, the study team will contact you to offer your baby an extra dose of vaccine. When the study finishes, you and your baby will continue to be looked after by your GP.

**What if relevant new information becomes available?**
Sometimes we get new information about vaccines or vaccination schedules that might be relevant to this study. If that happens or if the study is stopped for any reason, we will write to you and your GP with information about you and your baby's continuing care.

**What if there is a problem?**

- **Complaints**

If you have concerns about any aspect of this study, you can contact the study organiser or the Chief Investigator (contact details below), who will do their best to answer your questions. If you remain unhappy and wish to complain formally, then you can do this through the NHS Complaints Procedure, details of which can be obtained at www.nhs.uk or by phoning 0845 601 3012

- **Harm**

We do not anticipate any harm resulting from obtaining a blood samples. All vaccines used in this study are licensed and covered by the Manufacturers' product liability. If something does go wrong and you or your child is harmed during the study because of someone's negligence, then you may have grounds for legal action for compensation against the PHE, but you may have to pay your legal costs. The National Health Service complaints mechanism will still be available to you.

**Will our participation in the study be kept confidential?**
Personal data may include name and address of you and your baby as well as the blood test results and relevant medical information that would allow us to analyse the results and answer the study objectives. The only people with access to this information will be employees of the PHE, SGUL, Department of Health or regulatory authorities who may wish to check the study is being carried out within the appropriate guidelines. The data will only be used for the purposes of this study, will be stored in secure PHE facilities and will be destroyed after an appropriate time period, which may be a number of years.

**Involvement of my GP**
With your permission, we will send a letter to you and your baby's GP to let them know that you are both taking part in the study. We will also inform your GP of your baby's blood results and all vaccinations so that they can be added to your baby's medical records.

**What will happen to any samples from me and my baby?**
The blood samples we take for this study will be labelled with a study number and tested anonymously in fully certified PHE laboratories. We will write to you and your GP with the results of

the antibody levels and, if needed, recommendations for further vaccination. Once the tests are complete, there may be small amounts of blood remaining from you and/or your baby that we will use to try and understand how the whooping cough vaccine in pregnancy protects babies. We would like your permission to use any blood remaining after these tests to help us better understand why some children develop infections and how vaccines work to protect children. You will be asked to consent for this separately in the consent form. If you are not happy for the left over blood sample to be used for any other tests, please cross out the relevant section in the consent form and the samples will be destroyed after antibody testing. Your decision regarding the leftover blood will not affect your or your baby's participation in the study.

**What will happen to the results of the research study?**

We plan to publish the results in a medical journal that will be accessible to the public. The results of the study will also be reported to the Department of Health to help planning national vaccination programmes in the future. None of the reports will contain any information that might allow the readers to identify any mother or baby who took part in the study. At the end of the study, we will also write to all participating families to summarise the overall findings

**Who is organising and funding the research?**
The study is funded by the UK Department of Health and is being organised by PHE and St George's University of London. PHE is part of the Department of Health and has the remit of protecting the health and well-being of the population.

**Who reviewed the study?**
All research in the NHS is looked at by an independent group of people, called a Research Ethics Committee to protect the safety, rights, well-being and dignity of individuals. This study has been reviewed and given favourable opinion by the **XXXXX** Research Ethics Committee (Ref: **XXXXX**). Details of this study can be found on the following website: www.ClinicalTrials.gov (Ref: **XXXXX**).

**Further information and contact details.**
If you have any questions, you can ask your study team or contact the organisers, Professor Elizabeth Miller on 020 8327 7430 or Professor Paul Heath on 020 8725 5980.

We do hope that you and your family will take part in this study. Your contribution could be an important step towards the continual improvement of vaccine policy in the UK.

You and your child(ren) may be invited to take part in future studies with us by virtue of the GP surgery or hospital that you attend. We reassure you that invitation for future studies will not be related to individual results we obtain from this study.

| |
|---|
| Sticker with iMAP2 study team name and contact details (depending on site) |

#### 1.8 Covering letter 2 for use at St George's Hospital in the postnatal period (CovLetSGH2)

Public Health England

St George's Healthcare NHS Trust

St George's University of London

Dear Parent,

**Re: Clinical study being carried out at St George's Hospital - immunising Mums Against Pertussis (iMAP2)**

St George's Hospital and St George's University are working with Public Health England (PHE) on a study to compare two whooping cough vaccines that are currently used in pregnancy. The study also includes women and their infants who have not received a whooping cough vaccine in pregnancy. PHE is part of the Department of Health and has the remit of protecting the health and well-being of the population.

We are giving you this letter as you have recently had a baby. If you did not receive a whooping cough vaccine in pregnancy we would like you to consider whether you would like to take part in the study, which will allow you to have your child's antibody levels checked and an extra dose of vaccine given where indicated. The enclosed leaflet gives a brief summary about the study and tells you how you can receive more information.

Please note that neither St George's University nor PHE has your name or address details and will only contact you if you request further information.

If you do not want a member of the study team to contact you, you need do nothing further and please ignore any reminder card that we may send you. This will not affect your or your baby's routine medical care.

Yours sincerely

Representative of Maternity Services, St George's Hospital: name and signature

#### 1.9 Covering letter 2 for use by GPs in South London for women in the postnatal period (CovLetSGH-GP2)

Dear Parent,

**Re: Clinical study - immunising Mums Against Pertussis (iMAP2)**

We are working with Public Health England (PHE) and St George's University on a study to compare two whooping cough vaccines that are currently used in pregnancy. The study includes women and their infants who have not received a whooping cough vaccine in pregnancy. PHE is part of the Department of Health and has the remit of protecting the health and well-being of the population.

We are giving you this letter as you have recently had a baby. If you did not receive a whooping cough vaccine in pregnancy we would like you to consider whether you would like to take part in the study, which will allow you to have your child's antibody levels checked and an extra dose of vaccine given where indicated. The enclosed leaflet gives a brief summary about the study and tells you how you can receive more information.

Please note that neither St George's University nor PHE has your name or address details and will only contact you if you request further information.

If you do not want a member of the study team to contact you, you need do nothing further and please ignore any reminder card that we may send you. This will not affect your or your baby's routine medical care.

Yours sincerely

GP/representative signature & name

#### 1.10 Covering letter 2 for use by GPs at PHE sites for women in the postnatal period (CovLetPHE2)

Public Health England

St George's University of London

Dear Parent,

**Re: Clinical study - immunising Mums Against Pertussis (iMAP2)**

We are working with Public Health England (PHE) and St George's University on a study to compare two whooping cough vaccines that are currently used in pregnancy. The study includes women and their infants who have not received a whooping cough vaccine in pregnancy. PHE is part of the Department of Health and has the remit of protecting the health and well-being of the population.

We are giving you this letter as you have recently had a baby. If you did not receive a whooping cough vaccine in pregnancy we would like you to consider whether you would like to take part in the study, which will allow you to have your child's antibody levels checked and an extra dose of vaccine given where indicated. The enclosed leaflet gives a brief summary about the study and tells you how you can receive more information.

Please note that the PHE does not have your name or address details and will only contact you if you request further information.

If you do not want a Vaccine Research Nurse based at our practice to contact you, you need do nothing further and please ignore any reminder card that we may send you. This will not affect your or your baby's routine medical care.

Yours sincerely

GP/representative signature & name

#### 1.11 Pre-information leaflet 2 for use at all sites in the postnatal period (PreInfo2)

Public Health England

St George's Healthcare NHS Trust

St George's University of London

**Immunising Mums Against Pertussis (iMAP2)**

- Public Health England (PHE) and St George's University are working with GP surgeries to compare two whooping cough vaccines given in pregnancy, on behalf of the Department of Health. **We are also inviting a group of women who did not receive a whooping cough vaccine in pregnancy to participate in the study.**
- We would collect small blood samples from you and your baby. These would allow us to see the level of protection against infections that your baby received through the placenta and also to see how your baby responds to their vaccines. We would give your baby all their routine vaccines.
- If your baby has low antibody levels against certain infections, we will arrange for them to have a booster dose of vaccine to make sure they are well protected.

**If you have not received a whooping cough vaccine in pregnancy and would like further information please contact us by either:**

(i) Completing the reply slip at the bottom of this leaflet and returning it in the envelope provided.
(ii) Texting iMAP2 plus your name and GP surgery to the mobile number XXX

Thank You

✂-----------------------------------------------------------------------------------------------------------------------

I would like to receive further information about this study and I am happy for the study team to contact me.

Name: ____________________________________________ Due Date:___________

Address:__________________________________________________________________

_____________________________ Postcode:___________________________

Tel No:_______________________ Mobile no.______________________

Best time to contact me: __________ GP Surgery I attend: ___________________

#### 1.12 Participant information leaflet 2 for use at all sites in the postnatal period (PIL2)

**Immunising Mums Against Pertussis (iMAP2)**

**Full title: A head-to-head randomised controlled trial comparing two pertussis-containing vaccines in pregnancy and vaccine responses in UK mothers and their infants**

Public Health England and St George's University are conducting a study on behalf of the Department of Health comparing two whooping cough (pertussis) vaccines used in pregnancy. The study includes women who received one of these vaccines and women who did not receive a whooping cough vaccine in pregnancy.

We will measure the level of protective antibody in babies' blood before and after the routine infant vaccines given at 2, 3 and 4 months of age and then again after the booster vaccines given at 12 months. We will compare the level of antibody in infants whose mothers received a whooping cough vaccine in pregnancy with infants whose mothers did not receive a whooping cough vaccine in pregnancy. We will also measure the level of protective antibody that you have one year after the birth of your infant.

You have been contacted as you have recently delivered a baby. We would like to invite you and your baby to take part in this study if you did not receive a whooping cough vaccine in pregnancy.

Before you decide if you would like you and your baby to join this study, you need to understand the reasons why this study is being done and what it would involve. Please take the time to read the following information carefully and talk to others about the study if you wish.

**In summary:**

- The study involves taking three blood samples from babies to check their level of protective antibody to the vaccinations given under the routine UK schedule. It also involves one blood test from you.
- If the blood test shows low antibody levels against Meningitis C, pneumococcal or Hib germs, the study team will contact you to recommend an extra dose of vaccine to make sure that your baby is protected against these infections in the long-term. We will also offer an extra dose of polio vaccine if your baby has a low response and if you are planning to visit certain parts of the world where the risk of infection is highest, your nurse will discuss this with you.

**What is the purpose of the study?**

In September 2012, the UK Department of Health introduced an immunisation programme for pregnant women with whooping cough vaccination at 28-38 weeks of pregnancy. This recommendation was made following a marked rise in whooping cough cases across all age groups, but particularly in young infants who are at increased risk of severe disease.

This study compares the level of protective antibody that crosses the placenta to babies whose mother received one of the whooping cough vaccines and babies whose mother did not receive a whooping cough vaccine. This level of protective antibody is measured when the baby is 2 months old, just before the primary infant vaccinations. Levels of protective antibody will be measured again at 5 months of age, after the first set of routine infant vaccines and once more at 13 months of age, after the booster doses of vaccine. This is done to ensure that all babies achieve adequate levels of protective antibody.

This study will be important both for you to be reassured that your infant has adequate protection as well as helping the Department of Health continue to ensure the best protection is offered to our population.

**Why am I and my baby being invited to take part?**
We are inviting women who have delivered a baby recently and who have not received a whooping cough vaccine to take part in this part of the study. Women who delivered a baby at St George's Hospital or who are registered with participating GP surgeries across Hertfordshire, Gloucestershire and South London are being asked if they would like to participate. We plan to enrol up to 200 mothers and their babies in the overall study, around 50 of these women will have not received a whooping cough vaccine in pregnancy.

**Do we have to take part?**
You can decide whether you would like to take part. If you agree you will both take part, you will be asked to sign a consent form. Participation is entirely voluntary and you can withdraw from the study at any time without giving a reason and this would not affect either your or your baby's routine care. If you do not wish to participate or if you withdraw during the study, you will both be offered your routine immunisations through the GP surgery. If a blood sample has already been taken, you will need to decide if you want the sample to be destroyed, in which case you will need to inform us in writing. If the sample has already been tested, we will send you and your GP the results and discuss any further vaccinations advised with you.

**What will happen to my baby if we take part?**
This study involves three blood samples from your baby: one before and one after the routine infant vaccines (these are the vaccines given at 2, 3 and 4 months of age) and one after the booster vaccines (these are given at 12 months of age). These blood tests will allow us to estimate the level of protective antibody your baby receives through the placenta and how well they produce their own antibody in response to their vaccines. The study involves one blood sample from you one year after the birth of your baby. This allows us to see how well protective antibody persists in the mother.

If you agree for you and your baby to take part, a member of the study team will arrange an appointment to meet you at your convenience to answer any further questions that you may have. We will ask you some questions about your health and your baby's health and take a small sample (less than 1 teaspoon) of blood. We will offer ways to reduce discomfort in your infant for these tests.

The study team will give your baby all of the vaccinations according to the routine national schedule (the vaccines that would otherwise be given at your GP surgery).

In South London all the study visits will take place at your home or at St George's Hospital. In Hertfordshire and Gloucester the Vaccine Research Nurse based will meet you at your GP surgery or at home as appropriate.

If the study team is unable to obtain a blood sample from your baby, you will be given the option to re-schedule the appointment or miss that blood test. You are also free to withdraw at any time. The appointment will last about 45 minutes.

We will let your GP know that you and your baby are taking part in the study. We will write to you and your GP with the results of the blood tests taken when your baby is 1 year old. If your baby does not develop enough antibody against the Meningitis C, pneumococcal or Hib germs, then the study team will contact you to offer your baby an extra dose of vaccine so that your baby is protected against both these infections in the long-term. We will also offer an extra dose of polio vaccine if your baby has a low response and if you are planning to visit certain parts of the world where the risk of infection is highest, your nurse will discuss this with you.

The diagram below summarises what will happen in the study - visits shown according to the baby's age

| | **2 months** | **3 months** | **4 months** | **5 months** | **12 months** | **13 months** |
|---|---|---|---|---|---|---|
| **Mother** | | | | | | Blood test |
| **Baby** | Routine infant vaccinations<br><br>Blood test | Routine infant vaccinations | Routine infant vaccinations | Blood test | Routine booster vaccinations | Routine booster vaccinations<br><br>Blood test |

Routine vaccinations are set out in the "Red Book" which you will have been given by your midwife. The schedule is available at: http://www.nhs.uk/Tools/Pages/NHSvaccinationplanner.aspx#close

**What are the alternatives?**
If you do not wish to take part in this study, then you do not need to do anything. Your GP surgery will contact you for the routine vaccinations at the appropriate times.

**What are the possible disadvantages and risks as well as benefits of taking part?**
This study will involve blood tests that are not part of routine care. However, your baby will have the advantage of being offered an extra dose of vaccine if antibodies against polio, Hib, pneumococcal or Meningitis C are found to be low. There is also a benefit to the community, as our results will be used to inform future vaccination policy by the Department of Health.

Both vaccines are licensed and approved for adults. Repevax is currently recommended and used in pregnant women in the UK and Boostrix is recommended and used in pregnant women in other countries. Although the vaccines are not licensed for pregnant women this is because pregnant women are routinely excluded from clinical trials, and not because of any specific safety concerns or evidence of harm in pregnancy. The JCVI which is the body that advises the Department of Health on vaccine matters has stated that "There is no evidence of risk to pregnancy or the infant with inactivated viral or bacterial vaccines or toxoids such as those against diphtheria, tetanus, polio and pertussis (whooping cough)".

**What are the side-effects when taking part?**
The blood test may be uncomfortable but will be performed by experienced nurses or doctors who will use measures to reduce this. We do not anticipate any other side-effects.

**What happens when the study stops?**
We will write to you with the results of your baby's blood tests. If your baby is found to have low levels of protective antibody against the polio, Meningitis C, pneumococcal or Hib germs, the study team will contact you to offer an extra dose of vaccine. When the study finishes, your baby will continue to be looked after by your GP.

**What if relevant new information becomes available?**
Sometimes we get new information about vaccines or vaccination schedules that might be relevant to this study. If that happens or if the study is stopped for any reason, we will write to you and your GP with information about you and your child's continuing care.

**What if there is a problem?**

- **Complaints**

If you have concerns about any aspect of this study, you can contact the study organiser or the Chief Investigator (contact details below), who will do their best to answer your questions. If you remain unhappy and wish to complain formally, then you can do this through the NHS Complaints Procedure, details of which can be obtained at www.nhs.uk or by phoning 0845 601 3012

- **Harm**

We do not anticipate any harm resulting from obtaining a blood samples. All vaccines used in this study are licensed and covered by the Manufacturers' product liability. If something does go wrong and you or your child is harmed during the study because of someone's negligence, then you may have grounds for legal action for compensation against the PHE, but you may have to pay your legal costs. The National Health Service complaints mechanism will still be available to you.

**Will our participation in the study be kept confidential?**
Personal data may include name and address of you and your baby as well as the blood test results and relevant medical information that would allow us to analyse the results and answer the study objectives. The only people with access to this information will be employees of the PHE, SGUL,Department of Health or regulatory authorities who may wish to check the study is being carried out within the appropriate guidelines. The data will only be used for the purposes of this study, will be stored in secure PHE facilities and will be destroyed after an appropriate time period, which may be a number of years.

**Involvement of my GP**
With your permission, we will send a letter to you and your baby's GP to let them know that you are both taking part in the study. We will also inform your GP of your baby's blood results and all vaccinations so that they can be added to your baby's medical records.

**What will happen to any samples from my baby?**
The blood samples we take for this study will be labelled with a study number and tested anonymously in fully certified PHE laboratories. We will write to you and your GP with the results of the antibody levels and, if needed, recommendations for further vaccination. Once the tests are complete, there may be small amounts of blood remaining from you and/or your baby that we will use to try and understand how the whooping cough vaccine in pregnancy protects babies. We would like your permission to use any blood remaining after these tests to help us better understand why some children develop infections and how vaccines work to protect children. You will be asked to consent for this separately in the consent form. If you are not happy for the left over blood sample to be used for any other tests, please cross out the relevant section in the consent form and the samples will be destroyed after antibody testing. Your decision regarding the leftover blood will not affect yours or your baby's participation in the study.

**What will happen to the results of the research study?**

We plan to publish the results in a medical journal that will be accessible to the public. The results of the study will also be reported to the Department of Health to help planning national vaccination programmes in the future. None of the reports will contain any information that might allow the readers to identify any mother or baby who took part in the study. At the end of the study, we will also write to all participating families to summarise the overall findings

**Who is organising and funding the research?**
The study is funded by the UK Department of Health and is being organised by PHE and St George's University of London. PHE is an independent body that protects the health and well-being of the population and plays a critical role in protecting people from infectious diseases

**Who reviewed the study?**
All research in the NHS is looked at by an independent group of people, called a Research Ethics Committee to protect the safety, rights, wellbeing and dignity of individuals. This study has been reviewed and given favourable opinion by the **XXXXX** Research Ethics Committee (Ref: **XXXXX**). Details of this study can be found on the following website: www.ClinicalTrials.gov (Ref: **XXXXX**).

**Further information and contact details.**
If you have any questions, you can ask your study team or contact the organisers, Professor Elizabeth Miller on 020 8327 7430 or Professor Paul Heath on 020 8725 5980.

We do hope that you and your family will take part in this study. Your contribution could be an important step towards the continual improvement of vaccine policy in the UK.

You and your child(ren) may be invited to take part in future studies with us by virtue of the GP surgery or hospital that you attend. We reassure you that invitation for future studies will not be related to individual results we obtain from this study.

| Sticker with iMAP2 study team name and contact details (depending on site) |
| --- |

#### 1.13 Study Reminder Letter

Dear Parent

**Immunising Mums Against Pertussis (iMAP2)**

You should have received a letter recently informing you about a whooping cough vaccination study being conducted by Public Health England and St George's University of London. We are writing to check whether you received that letter or not.

- If you did receive the letter and have decided not to take part, then please ignore this reminder
- If you didn't receive the letter and would like to know more about the study, then please contact the study team (details below) who will arrange a convenient time to talk to you by telephone or in person
- If you received the letter and are thinking about taking part but have some questions regarding the study, then please contact the study team (details below) who will arrange a convenient time to talk to you by telephone or in person
- If you received the letter and have already agreed to take part, then you do not need to do anything else - thank you.

Thank you for your time.

Contact details –

Add for each site

#### 1.14 Parent Consent Form – randomised women and their infants

Centre: St George's / Gloucestershire/ Hertfordshire

Study code: iMAP2

Participant Identification Number: SN label

Title of Study: A randomised controlled trial comparing two pertussis-containing vaccines in pregnancy and vaccine responses in UK mothers and their infants

| | Participants please Initial box: |
|---|---|
| I ______________________________ confirm that I have read and understood the Patient Information Leaflet 1, **Version X**, dated **XX**. I have had the opportunity to consider the information, ask questions and have had satisfactory answers. | |
| I understand that both my and my baby's participation is voluntary and that I am free to withdraw us from the study at any time without giving any reason, without either of our medical care or legal rights being affected. | |
| I understand that relevant sections of both my and my baby's medical notes and data collected during the study may be looked at by individuals from Public Health England, St George's University Hospital or from regulatory authorities. I give permission for these individuals to have access to these records. | |
| I agree to my and my baby's GP being informed of our participation in the study | |
| I consent to be randomised to receive one of the two available whooping cough (pertussis) vaccines and I understand that I will not be able to choose which of these I receive. | |
| I consent for the study team to give me my vaccine and my baby's vaccines and to have the blood tests as set out in the Patient Information Leaflet 1 | |
| **Optional**: I consent to any remaining blood from me or my baby(s) to be used within Public Health England or St George's University of London, after the sample is anonymised, to | |

| |
|---|
| improve the understanding of infection and vaccines. **[NB. You and your baby can still take part in the study if you do not give consent for this. If you would like us to destroy any remaining sample, please strike through and do NOT initial this section]** |
| _______________________________ ____/____/____<br>Mother's signature Date:<br><br>Mother's name(block capitals) _______________________________ |
| I have explained and discussed the study with the above person. I have answered all their questions regarding the study and I am satisfied that the above signature denotes their informed consent to take part in the study.<br><br>_______________________________ ____/____/____<br>Signature of research nurse/doctor: Date:<br>Name(block capitals) _______________________________ |

When completed, 1 for participant's family; 1 for researcher master file (original); 1 to be kept in the infant's medical notes at the GP practice

#### 1.15 Parent Consent Form – unvaccinated women and their infants

Centre: St George's / Gloucestershire/ Hertfordshire (select centre)

Study code: iMAP2

SN label

Participant Identification Number:

Title of Study: A randomised controlled trial comparing two pertussis-containing vaccines in pregnancy and vaccine responses in UK mothers and their infants

| | Participant's please Initial box: |
|---|---|
| I _____________________________ confirm that I have read and understood the Patient Information Leaflet 2, **Version X**, dated **XX**. I have had the opportunity to consider the information, ask questions and have had satisfactory answers. | |
| I understand that both my and my baby's participation is voluntary and that I am free to withdraw us from the study at any time without giving any reason, without either of our medical care or legal rights being affected. | |
| I understand that relevant sections of both my and my baby's medical notes and data collected during the study may be looked at by individuals from Public Health England, St George's University Hospital or from regulatory authorities. I give permission for these individuals to have access to these records. | |
| I agree to my and my baby's GP being informed of our participation in the study | |
| I consent for the study team to give my baby their vaccines and for us to have the blood tests as set out in the Patient Information Leaflet 2 | |
| **Optional**: I consent to any remaining blood from me or my baby(s) to be used within Public Health England or St George's University of London, after the sample is anonymised, to improve the understanding of infection and vaccines. **[NB. You and your baby (s) can still take part in the study if you do not give consent for this. If you would like us to destroy any remaining sample, please strike through and do NOT initial this section]** | |

| |
|---|
| ______________________________________________ ____/____/____<br>Mother's signature Date:<br><br>Mother's name(block capitals) _____________________________________ |
| I have explained and discussed the study with the above person. I have answered all their questions regarding the study and I am satisfied that the above signature denotes their informed consent to take part in the study.<br><br>_________________________________________________ ____/____/_____<br>Signature of research nurse/doctor: Date:<br>Name(block capitals) ______________________________________________________ |

When completed, 1 for participant's family; 1 for researcher master file (original); 1 to be kept in the infant's medical notes at the GP practice

### 2 Protocol approval forms

#### 2.1 GP surgeries

##### General Practitioner Protocol Approval Form

**Title of Project:** A randomised controlled trial comparing two pertussis-containing vaccines in pregnancy and vaccine responses in UK mothers and their infants

**Short title:** immunising Mums against Pertussis, iMAP2

I confirm, on behalf of the GPs at the Surgery named below, that the attached protocol has been read, and it is agreed that patients registered at this surgery may be approached to participate in the study.

The Surgery is willing to support the study by sending out invitation letters to potential participants that are registered with the surgery. Specific arrangements for this will be discussed and agreed with the study team.

The Surgery will allow members of the study team to carry out study procedures (including vaccination and taking blood samples) at the surgery, and have appropriate access to patient records of study participants.

The Surgery will kindly inform the study team of any GP/A&E visits or hospitalisations involving study participants during the study.

______________________________ ______/_____/____

Signature of Surgery representative Date

______________________________ ____________________________

Name Title

Surgery address/ stamp

#### 2.2 Investigators

### Investigator Protocol Approval Form

**Title of Project:** A randomised controlled trial comparing two pertussis-containing vaccines in pregnancy and vaccine responses in UK mothers and their infants

**Short title:** immunising Mums against Pertussis, iMAP2

I approve this protocol and undertake that the study will abide by the provisions set forth therein. I agree to comply with the international conference on harmonisation tripartite guideline on good clinical practice

_____________________________ ____ /____/____

Investigator Signature Date

___________________________ ______________

Investigator Name Title

ADDRESS:

_____________________________________________________

_____________________________________________________

_____________________________________________________

_____________________________________________________

### 3 Correspondence with GP

#### 3.1 Letter to GP notifying of participation in study: randomised women

Public Health England

St George's University of London

**immunising Mums Against Pertussis (iMAP2)**

**Full title: A randomised controlled trial comparing two pertussis-containing vaccines in pregnancy and vaccine responses in UK mothers and their infants**

Dear Doctor

Re : Mother ________________________________________ Date of Birth: ___ / ___ / ___

Baby: _________________________________ Date of Birth: ___ / ___ / ___

The above-named person has agreed to participate in a study to assess their immune response to pertussis vaccination in pregnancy and the subsequent response of their baby the routine infant vaccines.

The study team will randomise the participating woman to receive one of the two available pertussis vaccines. The vaccine will be administered by the study team and we will advise you of the receipt of this vaccine.

We will collect blood samples from the mother and the infant and administer the infant's routine vaccines according to the following schedule, where visits are according to the baby's age:

| Visit | Pregnancy | Birth | 2 months | 3 months | 4 months | 5 months | 12 months | 13 months |
|---|---|---|---|---|---|---|---|---|
| **Mother** | REPEVAX<br>Or<br>BOOSTRIX-IPV<br><br>Blood test | Blood test | | | | | | Blood test |
| **Baby** | | Blood test or cord blood sample | Routine infant vaccinations<br><br>Blood test | Routine infant vaccinations | Routine infant vaccinations | Blood test | Routine booster vaccinations | Routine booster vaccinations<br><br>Blood test |

The blood samples will be tested for antibody responses to the vaccines at PHE laboratories. Infants with antibody concentrations below those considered to confer adequate protection against Hib, pneumococcal or MenC will be offered extra dose(s) of vaccine(s) as appropriate in order to ensure adequate protection against these infections. We will also offer an extra dose of polio vaccine if the baby has a low response and if is known to be visiting polio endemic countries prior to the pre-school booster, when they will receive a further polio vaccine anyway.

A copy of the blood test results taken at 13 months of age and details of the vaccines administered will be forwarded to your practice to be included in the baby's medical and immunisation records. If you would like any further information on this study, please do not hesitate to contact the study team.

Prof Elizabeth Miller and Professor Paul Heath

Sticker with Vaccine Research

Nurse name and contact details

#### 3.2 Letter to GP notifying of participation in study: unvaccinated women

Public Health England

St George's University of London

**Immunising Mums Against Pertussis (iMAP2)**

**Full title: A randomised controlled trial comparing two pertussis-containing vaccines in pregnancy and vaccine responses in UK mothers and their infants**

Dear Doctor

Re : Mother __________________________________________ Date of Birth: ___ / ___ / ___

Baby: ___________________________________ Date of Birth: ___ / ___ / ___

The above-named person has agreed to participate in a study to assess their infant's immune response to routine infant vaccines, though they themselves did not receive pertussis vaccination in pregnancy.

We will collect blood samples from the mother and the infant and administer the infant's routine vaccines according to the following schedule, where visits are according to the baby's age:

| | **2 months** | **3 months** | **4 months** | **5 months** | **12 months** | **13 months** |
|---|---|---|---|---|---|---|
| **Mother** | | | | | | Blood test |
| **Baby** | Routine infant vaccinations<br><br>Blood test | Routine infant vaccinations | Routine infant vaccinations | Blood test | Routine booster vaccinations | Routine booster vaccinations<br><br>Blood test |

The blood samples will be tested for antibody responses to the vaccines at PHE laboratories. Infants with antibody concentrations below those considered to confer adequate protection against Hib, pneumococcal or MenC will be offered extra dose(s) of vaccine(s) as appropriate in order to ensure adequate protection against these infections. We will also offer an extra dose of polio vaccine if the baby has a low response and if is known to be visiting polio endemic countries prior to the pre-school booster, when they will receive a further polio vaccine anyway.

A copy of the blood test results taken at 13 months of age and details of the vaccines administered will be forwarded to your practice to be included the baby's medical and immunisation records. If you would like any further information on this study, please do not hesitate to contact the study team.

Prof Elizabeth Miller and Professor Paul Heath

Sticker with Vaccine Research Nurse name and contact details

#### 3.3 Letter to GP notifying of vaccinations given (St George's site only – Hertfordshire and Gloucestershire nurses based in GP surgeries will note vaccinations directly to patient notes on computer systems)

**Immunising Mums Against Pertussis (iMAP2)**

**Full title: A randomised controlled trial comparing two pertussis-containing vaccines in pregnancy and vaccine responses in UK mothers and their infants**

Dear Doctor___________________,

Re: (Patient name)____________________________________________________________

Date of Birth: _______________________ Subject No. ____________________

NHS Number: _______________________

Further to our previous letter, the above-named infant has now completed the iMAP2 study and received the following vaccines.

| | Date | Name of vaccine | Batch number |
|---|---|---|---|
| 1. | __________________ | ____________________________ | ______________ |
| 2. | __________________ | ____________________________ | ______________ |
| 3. | __________________ | ____________________________ | ______________ |
| 4. | __________________ | ____________________________ | ______________ |
| 5 | __________________ | ____________________________ | ______________ |
| 6. | __________________ | ____________________________ | ______________ |
| 7. | __________________ | ____________________________ | ______________ |
| 8 | __________________ | ____________________________ | ______________ |
| 9. | __________________ | ____________________________ | ______________ |
| 10. | __________________ | ____________________________ | ______________ |

These vaccines will count as though given by your practice for target purposes and a record of the baby's vaccination has been forwarded to the Child Health Database.

If antibody levels to Hib, pneumococcal or meningococcal C antigens are found to be low at 13 months of age, we will inform the parents and ask them if they would like booster doses of the relevant vaccines to be given. If they do, we would appreciate it if these vaccines could be given in the surgery. . We will also advise an extra dose of polio vaccine if the baby has a low response and if is known to be visiting polio endemic countries prior to the pre-school booster, when they will receive a further polio vaccine anyway

Should the infant present to your surgery with any problem, or if you would like any more information, please do not hesitate to contact us.

Yours sincerely,

[PI: INDIVIDUALISE for each centre]

### 4.0 ADVERSE EVENTS

Definition of serious adverse events

An **adverse event** (AE) (or adverse experience) is any untoward medical occurrence in a subject to whom a medicinal product has been administered, including occurrences which are not necessarily caused by or related to that product.

An AE is considered as a **serious adverse event** (SAE) if the untoward medical occurrence:

- Results in death (in this study this will include mother, foetus or infant).
- Is life threatening (i.e., the subject was, in the opinion of the Investigator, at immediate risk of death from the event as it occurred); it does not refer to an event that hypothetically might have caused death if it were more severe.
- Requires or prolongs inpatient hospitalisation (more than 24 hours and not elective hospitalisation).
- Results in persistent or significant disability/incapacity (i.e., the event causes a substantial disruption of a person's ability to conduct normal life functions).
- Results in a congenital anomaly/birth defect.
- Requires intervention to prevent permanent impairment or damage.
- Is an important and significant medical event that may not be immediately life threatening or resulting in death or hospitalisation but, based upon appropriate medical judgment, may jeopardize the patient/subject or may require intervention to prevent one of the other outcomes listed above.

**Severity**

The severity of events will be determined by the Investigator as:

Mild: transient with no limitation in normal daily activity.

Moderate: some limitation in normal daily activity.

Severe: unable to perform normal daily activity.

**Relationship**

The relationship of the AE to the investigational product will be determined by the Investigator based on the following definitions:

- *Not Related*

The AE is not related if exposure to the investigational vaccine has not occurred, OR the occurrence of the AE is not reasonably related in time, OR the AE is considered unlikely to be related to use of the investigational vaccine, i.e. there are no facts (evidence) or arguments to suggest a causal relationship.

- *Unknown*

The administration of the investigational vaccine and AE may be considered reasonably related in time, BUT the AE could be explained by causes other than exposure to the investigational vaccine.

- *Related*

Exposure to the investigational vaccine and AE are reasonably related in time AND the investigational vaccine is more likely than other causes to be responsible for the AE, OR is the most likely cause of the AE.

**Adverse drug reactions**

An **adverse drug reaction (ADR)** is any untoward and unintended response in a subject to an investigational medicinal product (IMP) which is related to any dose administered to that subject.

An **unexpected adverse reaction** is an ADR the nature and severity of which is not consistent with the information about the IMP in the summary of product characteristics (or in the investigator's brochure if the IMP does not have a marketing authorisation).

**Reporting to regulatory bodies**

When an ADR is considered to be both unexpected and serious (i.e. a **suspected unexpected serious adverse reaction, SUSAR**), this must be reported to specified regulatory bodies (under current regulations at the time of preparing this protocol).

Throughout the study, adverse event reporting will be carried out in compliance with regulations that are applicable as and when they occur.

### 5 Participating Organisations and Study Personnel

**Immunisation, Hepatitis & Blood Safety Department, Public Health England, Colindale**

Professor Elizabeth Miller, Chief Investigator

Dr Shamez Ladhani, Dr Jo Southern, Dr Gayatri Amirthalingam, Co-investigators

Dr Nick Andrews, Statistical Advisor

Pauline Kaye, Data Manager

Teresa Gibbs & Deborah Cohen, Clinical Trials Administrators

Vaccine Research Nurses, Public Health England in Hertfordshire and Gloucestershire

**St George's, University of London**

Professor Paul Heath, Site Principal Investigator

Dr Chrissie Jones, Co-investigator

Dr Asma Khalil, Co-investigator

Research Nurses and midwives, St George's University of London

#### Participating laboratories

##### Laboratory evaluation of meningococcal and hepatitis B antibody responses by ELISA and meningococcal bactericidal assays

Prof Ray Borrow, Head - Vaccine Evaluation Consortium Laboratory, Manchester Medical Microbiology Partnership, Public Health England, Clinical Sciences Building, Manchester Royal Infirmary, Manchester M13 9WZ. Tel: 0161 276 8850. Fax: 0161 276 6792.

##### Laboratory evaluation of immune responses to diphtheria, tetanus, pertussis and Hib antigens by ELISA

Dr Bassam Hallis, General Project Manager, Public Health England, Centre for Emergency Preparedness and Response, Salisbury SP4 0JG. Tel 01980 612310. Fax 01980 611310.

##### Laboratory evaluation of pneumococcal antibody

Prof David Goldblatt, Immunobiology Unit, Institute of Child Health, 30 Guilford St

##### Laboratory evaluation of polio antibody

**Dr Gill Cooper,**
Division of Virology
NIBSC
Blanche Lane, South Mimms, Potters Bar.
EN4 3QG
