## Additional file 2 for "A phase IV, multi-centre, randomized clinical trial comparing two pertussis-containing vaccines in pregnant women in England and vaccine responses in their infants"

### Statistical Analysis Plan for the iMAP2 trial

**Version3**

**5th May 2017**

#### 1. Introduction

##### 1.1 Trial details

This is the statistical analysis plan for the following trial:

***A randomised controlled trial comparing two pertussis-containing vaccines in pregnancy and vaccine responses in UK mothers and their infants (immunising Mums Against Pertussis, iMAP2)***

The research question is to assess antibody responses following primary immunisation in children born to mothers vaccinated in pregnancy with one of two pertussis containing vaccines (by randomisation) and also in a smaller group of children born to unvaccinated mothers. The full rationale is given in the Protocol.

##### 1.2 Person responsible for the statistical analysis:

Nick Andrews, Statistics Unit, Public Health England. The analysis may be performed under supervision of the responsible statistician.

##### 1.3 Changes to this analysis plan:

A final version of this plan will be produced prior to provision of data to the statistician. Any additional analysis or deviation(s) from this analysis plan will be documented and the reasons given.

#### 2. Objectives

##### 2.1 Primary

PO: To compare anti-pertussis toxin (PT) IgG responses following primary immunisation with an acellular pertussis- containing vaccine in infants born to mothers who received REPEVAX in pregnancy compared to infants whose mothers received BOOSTRIX-IPV in pregnancy.

##### 2.2 Secondary

***Infant***

**SI1:** To compare antibody responses to pertussis antigens (concentration of IgG antibody to PT, pertactin (PRN), filamentous haemagglutinnin (FHA) and fimbrial antigens 2 and 3 (FIM 2 and 3]), tetanus toxoid, diphtheria toxoid and polio serotypes 1, 2 and 3 at birth amongst infants born to mothers who received REPEVAX in pregnancy compared to infants whose mothers received BOOSTRIX-IPV in pregnancy

**SI2:** To compare antibody responses to pertussis antigens [IgG to PT, PRN, FHA and FIM 2 and 3], Hib antigen [PRP], tetanus toxoid, diphtheria toxoid and polio serotypes 1, 2 and 3; meningococcal serogroup C serum bactericidal antibody titres and meningococcal serogroup C-specific IgG concentrations; 13 serotype-specific pneumococcal IgG concentrations and functional pneumococcal antibody studies at 2, 5 and 13 months of age (just before and one month after primary immunization and one month after booster vaccines) in infants born to mothers who received REPEVAX in pregnancy compared to infants whose mothers received BOOSTRIX-IPV in pregnancy and compared to infants whose mothers who did not receive pertussis vaccination in pregnancy

***Mother***

**SM1:** To determine concentrations of antibodies to pertussis antigens [IgG to PT, PRN, FHA and FIM 2 and 3] and tetanus toxoid, diphtheria toxoid and polio serotypes 1, 2 and 3, in pregnant women prior to administration of the REPEVAX or BOOSTRIX-IPV vaccine and at the time of delivery

**SM2:** To estimate the placental transfer ratio of antibodies to pertussis antigens [IgG to PT, PRN, FHA and FIM 2 and 3] and tetanus toxoid, diphtheria toxoid and polio serotypes 1, 2 and 3, in mothers who received REPEVAX in pregnancy compared to mothers who received BOOSTRIX-IPV in pregnancy

**SM3:** To compare concentrations of antibodies to pertussis antigens [IgG to PT, PRN, FHA and FIM 2 and 3] and polio serotypes 1, 2 and 3 at 12-13 months post-delivery in women who received REPEVAX or BOOSTRIX-IPV vaccine and to those women who did not receive pertussis vaccination in pregnancy

#### 3. Design

Full details are in the study protocol. Briefly women will be recruited at two sites during routine antenatal appointments and randomised to receive either REPEVAX or BOOSTRIX in pregnancy. A separate group of women who did not receive vaccination in pregnancy will be recruited at post natal visits as a control group.

##### 3.1 Study schedule

| Visit | V1 Pregnancy | V2 Birth | V3 2 months | V4 3 months | V5 4 months | V6 5 months | V7 12 months | V8 13 months |
|---|---|---|---|---|---|---|---|---|
| Visit Window | 28-32 weeks | 0-7 days hours after birth | 49-84 days of age | 21-42 days after visit 3 | 21-42 days after visit 4 | 21-42 days after visit5 | 353-390 days of age | 21-42 days after visit 7 |
| **Vaccinated mothers:** | REPEVAX/BOOSTRIX-IPV Blood MA | Blood MB | | | | | | Blood MC |
| **Unvaccinated Mothers** | | | | | | | | **Blood MC** |
| **Infants born to vaccinated mothers:** | | Blood BA | Routine* Blood BB | Routine* | Routine* | Blood BC | Routine* | (MMR**) Blood BD |
| **Infants born to unvaccinated mothers:** | | | Routine* Blood BB | Routine* | Routine* | Blood BC | Routine* | (MMR**) Blood BD |

*routine schedule is 2m: Infanrix/Prev13/Bexsero/Rotarix, 3m: Infanrix/MenC (Neis)/Rotarix, 4m: Infanrix/Prev/Bexsero, 12m: Menitorix/Prev/Bexsero/MMR. **MMR can be given at 12m or at 13m.

##### 3.2 Randomisation and blinding

Randomisation is from a block randomisation list produced prior to the study. On recruitment to the study, each subject will be allocated, in order of inclusion, the next available subject number. The study number will define the group to which the mother is assigned and which pertussis-containing vaccine she will receive. Mothers and their infants who are recruited in the postnatal period as part of the control group will not be randomised. Laboratory staff that test maternal and infant blood samples for vaccine responses will be blinded to the group allocation. Pregnant women and other study personnel will not be blinded. Though written informed consent will be taken at the first visit, continued consent to participate will be ascertained at each subsequent visit.

**4. Sample Size**

Based on sample size calculations below, it is proposed that a convenience sample of 65 (+/-10) mother-infant pairs will be recruited into each of the two groups that received a pertussis vaccine in pregnancy, with a control group of 30 (+/-10) women and their infants. Up to 200 mother-infant pairs will be recruited to take account of mother-infant pairs lost to follow-up.

The sample size calculation is based on data from the recent P13UK study performed by PHE. Based on these results, the standard deviation of the post-primary vaccination GMCs (log-10 scale) was:  PT = 0.28, Hib =0.76, MenC SBA = 0.50, Tet =0.37.  Based on these SD with 65 per study arm fold differences of 1.38,2.37,1.77 and 1.52 would be detectable between the vaccine arms for PT, Hib, MenC and Tet respectively.  In terms of percentages reaching putative  protective levels 95% in one arm would be different to percentages below 77% in the other arm.  The protocol has more details on other sample sizes.

**5. Increasing the size of the unvaccinated group**

Due to the small size of the unvaccinated arm (control group), this will be increased by including results from babies of unvaccinated mothers who participated in the Infanrix Hexa study (18 babies consenting from 2014-January 2016). These results are from testing in the same Laboratory. The schedule for infants in this study was

2m: Infanrix-Hexa™ &PCV13 & Rotarix™
3m: Infanrix-Hexa™ & Menitorix or NeisVacc™ & Rotarix™
4m: Infanrix-Hexa™ & PCV13
12m: PCV13 & MMR & Menitorix™

With bloods at 4m,5m,12m,13m
Children will also have received Bexsero (at 2,4m or in a 1 or 2 dose catch up) if born from May 2015.

Therefore vaccines are the same other than hepB added as Infanrix-hexa and MenC given at 3m. For the control group Meningococcal outcomes will therefore not be assessed. The results of the other anitbodies (pertussis, dip, tet, Hib, polio and pneumo) will be compared first between these 18 and the controls recruited in this study and only combined if differences are not significant at $p<0.005$ to allow for multiple comparisons).

**6. End points**

Shorthand for antibody measures:

- IgG to Pertussis antigens: **PT, PRN, FHA, FIM2, FIM3**

- Polio Titre to serotypes 1,2,3: **Polio1, Polio2,Polio3** (also **PolioX_8** for ≥8)
- Anti-tetanus toxoid IgG: **TT** (Also **TT0.10 TT1.00** mean conc≥0.10 and ≥1.00)
- Anti-diphtheria toxoid IgG: **Dip** (Also **Dip0.10 Dip1.00** mean conc≥0.10 and ≥1.00)
- Anti-PRP IgG: **Hib** (Also **Hib0.15 Hib1.00** mean conc≥0.15 and ≥1.00)
- Meningococcal serogroup C serum bactericidal antibody titre : **SBA** (Also **SBA8**, **SBA128** for ≥8 and≥128)
- meningococcal serogroup C-specific IgG: **MenC**
- Serotype-specific pneumococcal IgG for 13 vaccine serotypes: **Pn_x** (where x is the serotype).
- Functional pneumococcal antibody level: **Pn_fun_x** (where x is the serotype)

The end points are labelled to match the objectives.

##### 6.1 Primary end point

**PE:** Fold change in PT from blood taken at 2 to 5 months (post primary vaccination) in infants of vaccinated mothers.

##### 6.2 Secondary end points

**SI1:** In the infant at birth (just vaccinated mothers group) - PT, PRN, FHA, FIM2, FIM3,TT (TT0.10/TT1.00) ,Dip (Dip0.10/Dip1.00).

**SI2:** In infants at 2,5 and 13 months (all groups) PT, PRN, FHA , FIM2, FIM3, Hib (Hib0.15, Hib1.00),TT (TT0.10/TT1.00) ,Dip (Dip0.10/Dip1.00), SBA (SBA8, SBA128), MenC, Polio1,Polio2,Polio3 (PolioX_8), Pn_x, Pn_fun_x.

**SM1:** In Mothers pre vaccination and at delivery (just vaccinated mothers group) - PT, PRN, FHA, FIM2, FIM3, TT (TT0.10/TT1.00) ,Dip (Dip0.10/Dip1.00).

**SM2:** In Mothers/Babies (cord blood) at delivery (just vaccinated mothers group) ratio of Baby to Mother PT, PRN, FHA, FIM2, FIM3, TT (TT0.10/TT1.00) ,Dip (Dip0.10/Dip1.00).

**SM3:** In Mothers (all groups) at 13 months: PT, PRN, FHA, FIM2, FIM3. (And for vaccinated mothers ratio with levels at delivery).

##### 6.3. Safety data

Diaries are not being collected. End points will be Any serious adverse event following a vaccine dose (yes/no) and Hospital visit related to a vaccination (yes/no).

#### 7. Analysis sets and dealing with missing values and censored data

##### 7.1 Analysis sets

Analysis will be as treated for safety data and by modified ITT for immunogenicity data. mITT means an individual must have blood samples with results for inclusion, but will be included irrespective of interventions received. A per protocol analysis will also be performed if more than 10% of individuals would differ between mITT and per protocol.

Major protocol violations will be documented. These include incorrect vaccine given, vaccines not given and vaccines or blood samples given out of the visit windows.

##### 7.2 Dealing with missing values and censored data

The reason for missing data will be indicated but missing data will not be imputed. Censored data will be given a value half the lower limit and twice the upper limit of the assay.

#### 8. Supply of data and locking of the database

Data will be entered into a database at PHE immunisation dept. This will be transferred to the Statistician through exports to Excel or as ASCII files. The data will then be imported into Stata for analysis. Checks will be performed to ensure correct transfer and the statistician will also perform logical checks of the data. Once satisfactory the database will be locked.

#### 9. Interim analysis

No formal Interim analysis is planned.

#### 10. Statistical Methods

##### 10.1 Statistical package

Stata version 13 or higher will be used.

##### 10.2 Significance level and confidence intervals

95% confidence intervals will be reported. Where comparisons are made between groups a 5% significance level will be used.

##### 10.3 Descriptive analysis

A description of number in the trial and with results at each stage will be produced in a flow chart (consort diagram). Descriptive tables of numbers by age (mother), sex (baby), ethnic group, study site, birth weight, gestational age, gestational age at vaccination, weeks prior to birth of vaccination, timing of blood at birth, receipt of influenza vaccination, breast feeding for first 2 months, and other baseline variables will be produced.

##### 10.4 Immunogenicity analysis

The end points in PE,SI1,SI2,SM1,SM2,SM3 are either concentrations, titres, ratios of concentrations or tires or binary values(for being over a threshold). For concentrations and titres (and ratios of these) results will be log transformed and the groups (REPEVAX vs BOOSTRIX-IPV, OR REPEVAX vs BOOSTRIX-IPV vs Control) compared by normal errors regression (equivalent to ANOVA for 3 groups or an unpaired t-test for 2 groups). Geometric means for each group will be calculated with 95% confidence intervals and Geometric mean fold ratios between groups with 95% CIs. If the log transformed data are not normally distributed then the kruskal-wallis test will be used to compare the groups.

Proportions above titre thresholds will be compared using Fisher's exact test and proportions calculated within groups with 95% exact confidence intervals.

For persistence (SM3) geometric mean fold changes will be calculated with 95% CI.

##### 10.5. Additional exploratory analyses

In regression analyses of titres the effect of titre pre vaccination (at 2m) on post primary responses (at 5m and also 13m to the same and different antigens) will be assessed. Regression models to allow for timing of the blood after primary vaccination (or birth), sex, ethnicity, gestational age at vaccination (or weeks prior to birth), study site, gestational age and birth weight will be fitted to investigate these factors and adjust if necessary.

Prior receipt of pertussis vaccination in pregnancy will be investigated for the pre-vaccination maternal blood.

The association between antibodies at birth in infants and at 2 months will be investigated.

The association between breast feeding and post primary responses.

##### 10.6 Safety analysis

The proportion with SAE and Hospital visit will be calculated for each group.

#### 11. Reports

The results of the analysis will be written up in a statistical analysis report. This report can then be used to assist in the production of papers for publication and the main study report. It may also be included as an addendum to the main report. The statistician who analysed the data should see any further reports based on the statistical analysis report.
